# Amyloid PET predicts atrophy in older adults without dementia: Results from the AMYPAD Prognostic & Natural History Study

**DOI:** 10.1101/2025.04.09.25324824

**Authors:** Leonard Pieperhoff, Luigi Lorenzini, Sophie Mastenbroek, Mario Tranfa, Mahnaz Shekari, Alle Meije Wink, Robin Wolz, Sylke Grootoonk, Craig Ritchie, Mercè Boada, Marta Marquié, Wiesje van der Flier, Rik Vandenberghe, Bernard J. Hanseeuw, Pablo Martínez-Lage, Pierre Payoux, Pieter Jelle Visser, Michael Schöll, Giovanni B. Frisoni, Andrew Stephens, Christopher Buckley, Gill Farrar, Frank Jessen, Oriol Grau-Rivera, Juan Domingo Gispert, David Vállez García, Henk Mutsaerts, Frederik Barkhof, Lyduine E. Collij, the AMYPAD consortium

## Abstract

The impact of amyloid-β (Aβ) accumulation on regional brain atrophy in preclinical Alzheimer’s disease (AD), and its interaction with risk factors like sex and *APOE*-ε4 carriership, remains unclear. In this study, we examined these associations in a population of older adults without dementia and evaluated the potential of Aβ-PET for risk stratification.

We included 1329 participants (56% female) with an age of 68.2 ± 8.78 years from the prospective multi-center AMYPAD Prognostic and Natural History Study who underwent [^18^F]Flutemetamol or [^18^F]Florbetaben Aβ-PET and T1-weighted MRI, with longitudinal data for 684 participants (median follow-up=3.4 years). Linear mixed models assessed the effect of baseline Aβ burden through the Centiloid approach on longitudinal changes in regional gray matter volume and thickness. Sensitivity analyses were performed in cognitively normal only (CDR=0) individuals and while correcting for CSF p-tau181 and t-tau. A second model investigated the effects of sex or APOE-ε4 carriership.

Baseline global Aβ was predictive of widespread atrophy in several brain regions, most strongly in the fusiform (β_Volume_=-0.006, β_Thickness_=-0.009), hippocampus (β_Volume_=-0.005), posterior cingulate (β_Volume_=-0.006), and precuneus (β_Volume_=-0.004, β_Thickness_=-0.007), also when investigating only in cognitively normal individuals. Only fusiform atrophy (β_p-tau_=- 0.011; β_t-tau_=-0.011) remained predicted by Aβ when correcting for p-tau181 or t-tau. Temporal atrophy was exacerbated in women, while frontal, lateral-temporal and hippocampal atrophy was exacerbated by carriership of at least one *APOE-*ε4 allele. Aβ burden had a larger impact on changes in thickness than volume.

Our findings suggest that in older adults with mostly preserved cognition, baseline Aβ-PET predicts future brain atrophy, with fusiform atrophy showing independence from tau pathology and Aβ-dependent atrophy being exacerbated in region-dependent manners in females and *APOE*-ε4 carriers. Regional cortical thickness may serve as a sensitive marker for early Aβ-related neurodegeneration and aid in stratifying risk in AD prevention trials.

## 1. Introduction

Brain atrophy is a key feature of neurodegenerative diseases and a common surrogate endpoint in Alzheimer’s disease (AD) clinical trials (Schwarz et al., 2019). In its clinical stage, AD-associated atrophy and cognitive decline are closely linked to tau deposition (Bejanin et al., 2017). However, as the field of anti-amyloid therapy is shifting towards early intervention, there is a need to better understand the predictive value of early amyloid-beta (Aβ) accumulation on atrophy in the preclinical stages of the disease.

AD-associated atrophy has commonly been described first in medial-temporal areas, followed by more widespread involvement of the neocortex (Chételat et al., 2012; La Joie et al., 2020; Tosun et al., 2011). Evidence on the effect that early amyloid deposition exerts on regional atrophy are conflicting, however, with some highlighting frontoparietal rather than medial-temporal atrophy patterns (Martikainen et al., 2019; Mattsson et al., 2014), while others have reported no observed atrophy (Josephs et al., 2008) or even increased volumes (Chételat et al., 2010; Ingala et al., 2021). This conflicting evidence may be partially explained by the heterogeneity of the measures used to quantify atrophy, e.g. volume or thickness, and local, regional and global analyses as well as the investigated disease stages. In a recent study, CSF measures of amyloid-beta 42 were predictive of medial-temporal atrophy rates in individuals without dementia, suggesting that amyloid deposition may lead to atrophy independently of CSF p-tau (Cacciaglia et al., 2025). However, CSF shows an early plateau effect and is less effective in tracking the extent of amyloid pathology compared to amyloid positron emission tomography (PET), which can be substantial even in cognitively unimpaired individuals (Collij et al., 2021; Salvadó et al., 2019). Taken together, these results underscore the need to further characterize the possible relationship between PET measured cerebral amyloid deposition and neurodegenerative processes in the earliest stages of the disease, with the goal of informing early secondary or even primary prevention strategies.

In addition, the individual vulnerability to AD-related neurodegenerative processes due to demographic and genetic risk factors must be taken into account,(Kepp et al., 2023) Previous findings regarding the effect of *APOE-*ε4 carriership have reported either increased volume loss in lateral-temporal and superior-frontal regions in a cohort of amyloid-positive subjects with mild cognitive impairment (MCI)(Sauty and Durrleman, 2023) or widespread cortical thinning in prodromal AD *APOE*-ε4 non-carriers compared to carriers.(Mattsson et al., 2018) These discrepancies may have arisen from the isolated examination of gray matter volume and thickness, highlighting the necessity for concurrent analyses of these image-derived phenotypes (IDPs). Regarding sex differences, women show higher AD prevalence,(Gustavsson et al., 2023) faster accumulation of cortical tau as measured with PET(Wisch et al., 2021) and greater cortical atrophy(Sauty and Durrleman, 2023) compared to males in clinical stages. In contrast, women in preclinical cohorts show a delayed and slower atrophy rate despite greater tau burden,(Buckley et al., 2019; Sauty and Durrleman, 2023) illustrating the complexity of this relationship. Hence, understanding the moderating effects of demographic and genetic risk factors on the association between Aβ burden and subsequent atrophy is crucial for informing clinical practice and patient selection/stratification in clinical trials.

The current study aims to determine the influence of global cortical amyloid burden on regional brain atrophy. To this end, we examined regional changes in brain volume and thickness in a large cohort composed of older adults without dementia from the Amyloid Imaging to Prevent Alzheimer’s Disease (AMYPAD) Prognostic Natural History Study (PNHS). We further investigated how the relationship between Aβ and atrophy is influenced by sex and *APOE-*ε4 carriership, and the additive predictive value of utilizing longitudinal amyloid-PET data.

## 2. Materials and methods

### 2.1 Participants

Data for this study were drawn from the AMYPAD PNHS v202306 (*N*=1585).(García, 2023) The AMYPAD PNHS (EudraCT: <u>2018-002277-22</u>) is a multi-center cohort of non-demented participants to determine the value of amyloid PET in clinical- and research settings; the study design has been described in detail previously.(Lyduine E. Collij et al., 2022; Lopes Alves et al., 2020) Briefly, eligibility criteria for inclusion in the AMYPAD PNHS were no history of dementia (clinical dementia rating (CDR) < 1), age above 50 years, and being able to undergo an amyloid-PET and MRI scan. The studies were reviewed and approved by Medical Ethical Committee of the University Medical Center Amsterdam, location VUmc and all local sites. The studies were conducted according to the principles of the Helsinki Declaration of 1975, as revised in 2008, and all human participants gave written informed consent. Median time difference between the MRI and PET acquisition was 50 days (*IQR* 99 days). A subset of 684 participants underwent at least one follow-up PET and MRI, at a median follow-up of 3.4 years (± 1.4 years) after baseline. None of the study participants participated in clinical trials of anti-amyloid therapy throughout the duration of the study.

### 2.2 A**β**-PET Acquisition and Quantification

PET scans were acquired 90-110 minutes p.i. of 185 MBq (±10%) for [^18^F]Flutemetamol and 350 MBq (±20%) for [^18^F]Florbetaben, consisting of 4 frames of 5 minutes according to the standard protocol for each tracer.(Barthel et al., 2011; Curtis et al., 2015) Image analysis was performed centrally using IXICO’s automated workflow. Briefly, PET frames were co-registered, averaged, and aligned to the corresponding MRI scan, which was parcellated using a subject-specific multi-atlas approach, i.e. the learning embeddings for atlas propagation (LEAP) parcellation procedure.(Wolz et al., 2010) SUVr images were obtained using LEAP parcellation masks using the whole cerebellum as a reference region in native space. In order to pool Aβ-PET data across sites, SUVr values were transformed to Centiloids (CL) using the standard Global Alzheimer’s Association Interactive Network target region as a measure of global amyloid burden.(Klunk et al., 2015)

Participants were grouped in low (CL≤20), intermediate (20<CL≤40), and elevated (CL>40) Aβ stages, corresponding to cut-points in current early secondary prevention trials(Rafii et al., 2023) and based on derived cut-offs to define reliable worsening and detection of amyloid plaques on neuropathology.(Jack et al., 2017; Villeneuve et al., 2015)

### 2.3 MRI Acquisition and Quantification

3D T1-weighted MR images were acquired on Philips (n=1340), Siemens (n=761) or GE HealthCare (n=24) scanners, all of which had a field strength of 3T. All longitudinal MRI were acquired on the same scanner model. Cortical segmentation was performed with FreeSurfer v7.1.1 (<u>surfer.nmr.mgh.harvard.edu/</u>), including motion correction, skull-stripping, brain parcellation and estimation of regional gray matter volume and thickness. The details of these procedures are described elsewhere.(Reuter et al., 2012) The final IDPs included estimated total intracranial volume (eTIV), volumes of six sub- and allocortical regions of interest encompassing the lateral ventricles, thalamus, caudate, putamen, hippocampus and amygdala, as well as volume and thickness of 34 bilateral cortical regions of interest covering the whole cerebral cortex defined using the Desikan-Killiany atlas.(Desikan et al., 2006) The parcellations were visually inspected, resulting in the exclusion of 36 subjects due to segmentation errors. IDPs were harmonized using neuroCombat v1.0.13 to remove batch effects, while preserving the biological effects of age, sex and global CDR scores (**Supplementary** Fig. 1).(Fortin et al., 2018) Median follow-up time was 3.40 years (± 1.4 years) with a median of one follow-up visit.

### 2.4 Statistical analysis

All analyses were carried out in R v4.3.0 (www.r-project.org/). Differences in demographics between Aβ groups were assessed using ANOVA and Chi-Square tests for continuous- and categorical variables, respectively.

To assess the effect of baseline global cortical Aβ burden on subsequent regional neurodegeneration, linear mixed-effects models (LME) implemented via the R package “lme4” v1.1-35.3 with random slope and intercept were used. The main predictors included baseline amyloid burden (continuous CL), follow-up time, and their interaction while correcting for age and CDR at baseline, sex, eTIV, and their interaction; in R equation format: “GM Volume/Thickness ∼ 1 + Centiloid_Baseline_*Time + Age_Baseline_ + Sex + eTIV + CDR_Baseline_ + (1 + Time | Subject)”.(Dhamala et al., 2022). As a sensitivity analysis, these analyses were repeated in the CDR=0 subset (*N*=1099).

To investigate whether baseline amyloid is predictive of atrophy independently of baseline CSF p-tau181 and t-tau levels, analyses were repeated in a subset of participants with available CSF biomarkers (N=428; N=808 visits). First, the same base model was run, followed by including baseline t-tau or baseline p-tau181 as an additional covariate.

To assess the added value of longitudinal Aβ-PET measurements in predicting subsequent atrophy, we also repeated the main analyses in the subset of participants with longitudinal PET (*N*=684), Cross-sectional-only and longitudinal model fits were compared using the Akaike Information Criterion derived from ANOVA.

Finally, the effect of *APOE*-e4 carriership and sex on amyloid-atrophy relationships was assessed using one additional LME model with two added three-way interactions, namely amyloid*time\**APOE* and amyloid*time*sex.

All IDPs and CL values were z-scaled to obtain standardized regression coefficients. The significance level was set at α<0.05 after applying the Benjamini-Hochberg procedure to correct for a false discovery rate.(Benjamini and Hochberg, 1995)

## 3. Results

Baseline demographics can be found in **Table 1**. The majority of individuals were cognitively unimpaired (CDR=0:*N*=1070, 81%), with an overall average MMSE score of 28.82 (±1.54). Mean age was 68.18 (±8.78) years, 741 (56%) were female, and 117/260 (8.8%/19.6%) were considered to have intermediate and elevated Aβ, respectively. Carriership of at least one allele of *APOE*-ε4 was 40% overall, with stepwise increases in carriership in the amyloid groups with 300/58/159 (32%/51%/66%) *APOE*-ε4 carriers in low, elevated and elevated AB participants, respectively. Follow-up time did not differ across amyloid groups with an overall median follow-up time of 3.4 (MD=1.40) years, although overall availability of follow-up data differed between amyloid groups with more longitudinal data available for subjects with low Aβ (**Supplementary** Fig. 2). Individuals with elevated Aβ more frequently had a CDR=0.5, lower MMSE scores, and a higher frequency of *APOE-*ε4 carriership compared to low Aβ individuals. Only two individuals converted to CDR=1 at follow-up, both of which had elevated Aβ at baseline. Sex- and *APOE*-stratified demographic characterizations can be found in **Supplementary Table 1** and **Supplementary Table 2**, respectively. Male participants were older (68.81 years compared to 67.68 years), had higher educational level, and a larger proportion of very mild cognitive impairment (24% compared to 16%), but without any difference in MMSE or Aβ burden. People with one or more *APOE-* ε4 allele were younger (66.56 years compared to 69.02 years), had higher proportion of very mild cognitive impairment (23% compared to 16%), lower MMSE (28.67 compared to 28.99), and higher Aβ burden (29.57 CL compared to 11.33 CL).

**Table 1.** Demographics and Clinical Characteristics at Baseline.

|  | **Overall**<br>(n = 1,329)<sup>1</sup> | **Low Aβ**<br>(n = 952)<sup>1</sup> | **Intermediate Aβ**<br>(n = 117)<sup>1</sup> | **Elevated Aβ**<br>(n = 260)<sup>1</sup> | **p** |
| --- | --- | --- | --- | --- | --- |
| Age, years | 68.18 (8.78) | 66.47 (8.21) | 70.64 (8.30) | 73.33 (8.80) | <0.001<sup>2</sup> |
| Follow-Up, years | 3.74 (1.87) | 3.81 (1.88) | 3.54 (1.91) | 3.39 (1.75) | 0.037<sup>2</sup> |
| (Missing) | 585 | 355 | 58 | 172 |  |
| Sex, Female | 741 (56%) | 545 (57%) | 63 (54%) | 133 (51%) | 0.2<sup>3</sup> |
| Education, years | 14.56 (3.97) | 14.71 (3.96) | 14.47 (3.84) | 14.05 (4.01) | 0.075<sup>2</sup> |
| CDR |  |  |  |  | <0.001<sup>3</sup> |
| 0 - Normal | 1,070 (81%) | 833 (88%) | 90 (77%) | 147 (57%) |  |
| 0.5 - Very mild | 259 (19%) | 119 (13%) | 27 (23%) | 113 (43%) |  |
| CDR at Follow-Up |  |  |  |  | <0.001<sup>4</sup> |
| 0 - Normal | 572 (88%) | 486 (92%) | 40 (78%) | 46 (68%) |  |
| 0.5 - Very mild | 76 (12%) | 45 (8%) | 11 (22%) | 20 (29%) |  |
| 1 - Mild | 2 (0%) | 0 (0%) | 0 (0%) | 2 (3%) |  |
| (Missing) | 679 | 421 | 66 | 192 |  |
| MMSE | 28.82 (1.54) | 29.06 (1.27) | 28.68 (1.52) | 27.91 (2.08) | <0.001<sup>2</sup> |
| (Missing) | 108 | 61 | 8 | 39 |  |
| APOE-ε4 carriership (% carriers) | 517 (40%) | 300 (32%) | 58 (51%) | 159 (66%) | <0.001<sup>3</sup> |
| (Missing) | 40 | 17 | 3 | 20 |  |
| Amyloid PET, Centiloid | 19.51 (32.33) | 2.81 (8.37) | 28.55 (6.05) | 76.58 (27.48) | <0.001<sup>2</sup> |
| est. Total Intracranial Volume, cm<sup>3</sup> | 1477.71 (178.43) | 1476.68 (177.60) | 1477.94 (196.79) | 1481.38 (173.41) | 0.7<sup>2</sup> |

Abbreviations: Aβ, amyloid β; CDR, Clinical Dementia Rating; MMSE, Mini-Mental State Examination; PET, Positron Emission Tomography; Amyloid stages are defined based on a Centiloid value of under 20 for low, over 40 for elevated Aβ and a subsequent intermediate Aβ between 20 and 40 CL.
<sup>1</sup>Mean (SD); n / N (%)
<sup>2</sup>Kruskal-Wallis rank sum test
<sup>3</sup>Pearson's Chi-squared test
<sup>4</sup>Fisher's exact test

### 3.1 Effects of baseline A**β** burden on regional atrophy

LME results of the main models can be found in **Supplementary Table 3** and are visualized in **Figure 1**. Besides one significant positive effect of time on temporal pole volume, there was a significant negative main effect of time on all investigated cortical and subcortical gray matter volumes, suggesting widespread atrophy due to ageing-related neurodegeneration. This effect was most apparent in temporal and parietal regions as well as hippocampus and amygdala. Similar results were observed for cortical thickness, although posterior and anterior cingulate cortices showed thickening over time (**Figure 1A**). All effects remained significant with comparable effect sizes in the CDR=0 subset (**Figure 1D**).

**Figure 1.**
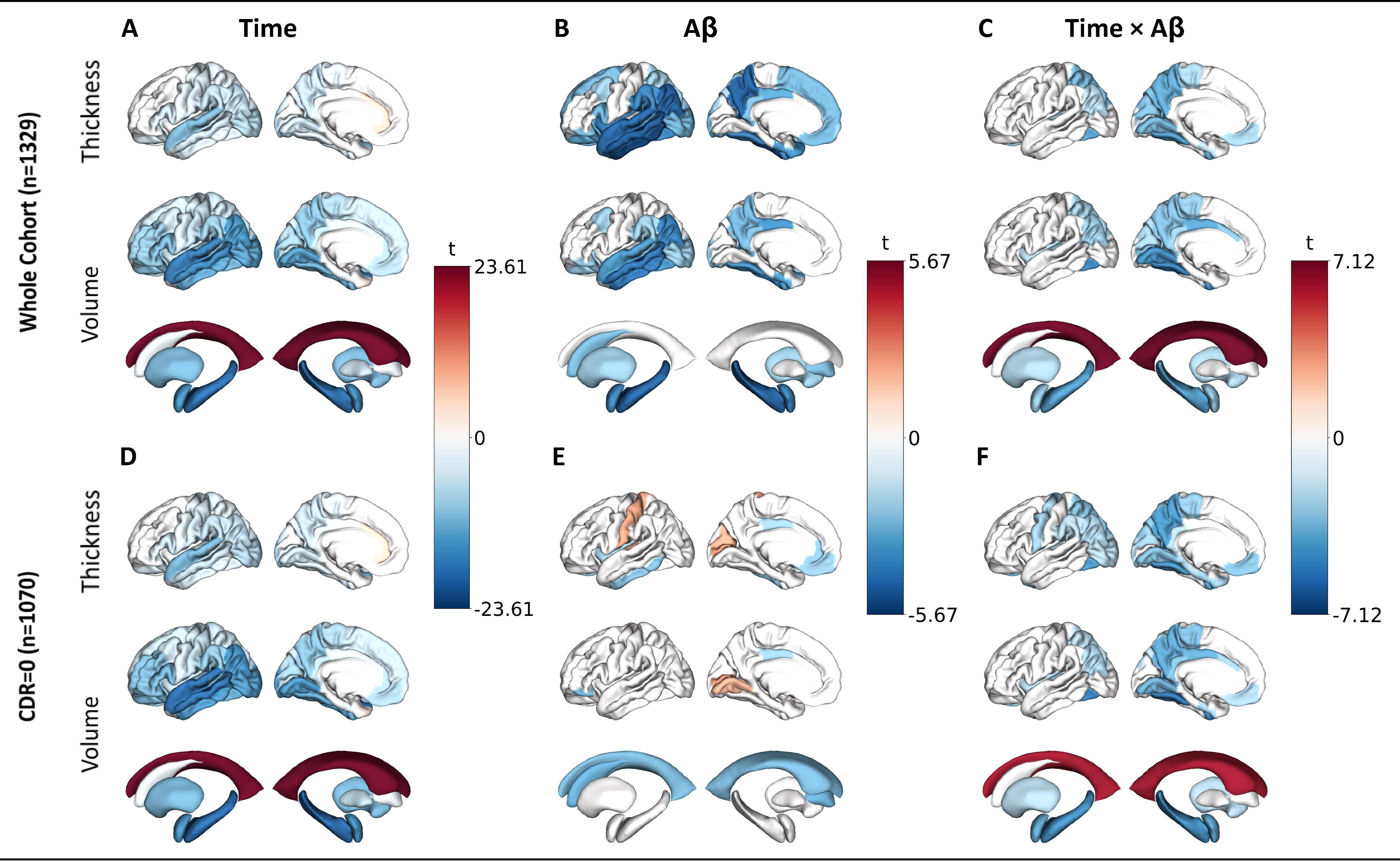
Main & interaction effects of time & amyloid at baseline on regional volumes & thicknesses, in all participants and cognitively unimpaired (CDR=0). Cortical mid-surface projections of FDR-thresholded LME main effects on cortical thickness, cortical volume and subcortical volume, of time (**A**) in all participants and (**D**) cognitively untimpaired (CDR=0), main effect of baseline cortical Aβ (**B**) in all participants and (**E**) cognitively unimpaired (CDR=0), and interaction effect of time and Aβ (**C**) in all participants and (**F**) cognitively unimpaired (CDR=0). Surface projections show the lateral and medial cortex respectively, representative of averaged bilateral effects. Aβ, amyloid β; CDR, Clinical Dementia Rating.

Higher baseline cortical amyloid burden was associated with lower baseline volumes at baseline in widespread AD signature regions, comprising lateral-temporal lobes, precuneus, posterior cingulate, amygdala, and hippocampus. These main effects and patterns also emerged for cortical thickness but were more distributed and stronger compared to volumes (**Figure 1B**). These effects were largely absent in the CDR=0 subset (**Figure 1E**).

Longitudinally (Centiloid*Time), higher baseline cortical amyloid burden was related to a widespread decrease of volume and thickness at follow-up across regions, with the strongest interaction effects observed in the posterior cingulate, fusiform and parahippocampal gyri (**Figure 1C**). Subcortically, this effect was most pronounced in the hippocampus and amygdala, followed by the putamen. Lateral ventricles showed progressive widening in relation to cortical amyloid burden. Except for hippocampal volume changes, most interaction effects were Aβ stage-dependent, with low and elevated Aβ groups showing significantly different slopes, but not low and intermediate Aβ groups. (**Supplementary** Fig. 3). In the CDR=0 subset, most effects remained significant with slightly lower effect sizes (**Figure 1F**).

In the CSF availability subset (N=428), fewer Centiloid*Time interactions remained significant after FDR-correction, albeit with the same patterns (Supplementary Table 4). When correcting for t-tau or p-tau181, only the fusiform volume atrophy remained significant (t-tau: β=-0.011, SE=0.003, p_FDR_=0.017; (p-tau: β=-0.011, SE=0.003, p_FDR_=0.018).

Stratifying for amyloid status rather than continuous Centiloids, faster fusiform atrophy rates were found in participants with elevated-compared to low Aβ burden (β=-0.054, SE=0.017, p_FDR_=0.007), but not for participants with intermediate-compared to low Aβ (β=-0.013, SE=0.018, p_FDR_=0.749; **Figure 2**).

**Figure 2.**
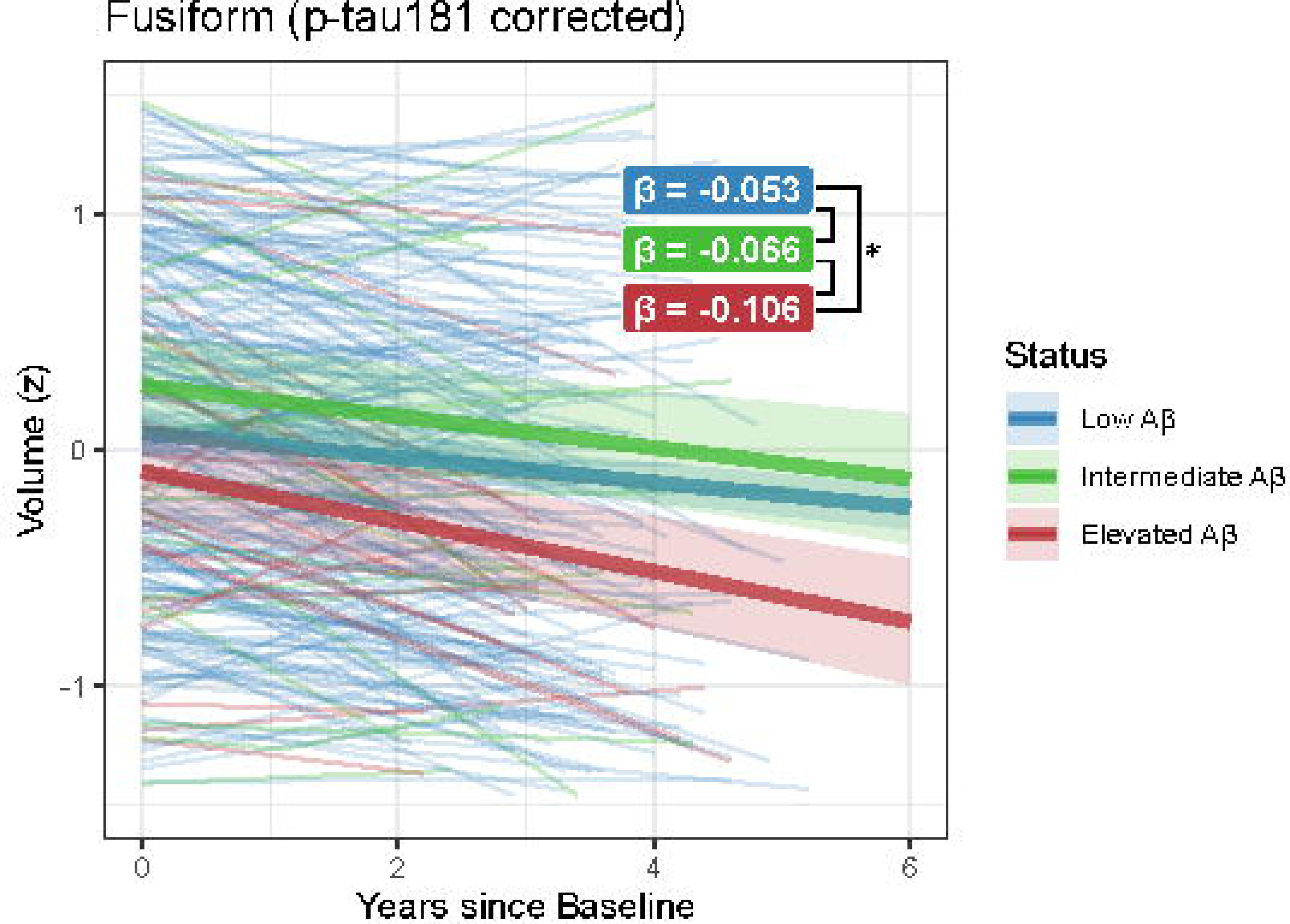
**Spaghetti plot of LME model effects of follow-up time on fusiform gyrus volume, stratified by amyloid status**. Fusiform volume (z-scaled, residuals after correcting for age, sex, intracranial volume, CDR and p-tau181) is plotted against time (in years) since baseline amyloid-PET and MRI, stratified in low (<20 CL), intermediate (20-40 CL) and elevated Aβ (>40 CL) groups. Fusiform atrophy is larger in participants with elevated Aβ compared to participants with low Aβ, indepent of p-tau181. Aβ, amyloid β; CDR, Clinical Dementia Rating; PET, Positron Emission Tomography.

### 3.2 Stratification by sex & *APOE-***ε**4 carriership

To investigate the effect of sex and *APOE-*ε4 carriership, the LME models were expanded with three-way interactions (i.e., amyloid*time*sex and amyloid*time\**APOE)*. Interaction effects between *APOE-*ε4 and time, sex and time, and *APOE-*ε4, sex and time are visualized in **supplementary Figure 4**. Over time, men had stronger loss of thickness in the superior temporal, inferior and superior parietal, and cuneus, as well as increased loss of volume in the caudate nucleus and faster increase of lateral ventricle size. Women only had stronger atrophy in the caudal anterior cingulate gyrus. *APOE-*ε4 carriers had decreased growth of lateral ventricle volume, and lower atrophy rates of entorhinal, temporal pole and frontal pole volume, as well as lower loss of frontal pole thickness. Effects of sex on the relationship between amyloid and brain atrophy were widespread (**Figure 3A**). Women showed a generally exacerbated effect of Aβ on atrophy compared to men, especially in lateral temporal regions and hippocampal volume (β=0.006), with relatively less Aβ-related atrophy only in the caudate nucleus (β=-0.007), pericalcarine volume (β=-0.006) and posterior cingulate thickness (β=-0.008). For *APOE-*ε4-carriers, more severe cortical thinning and loss of volume as a result of amyloid increase was observed compared to non-carriers (**Figure 3B**) especially in frontal and lateral temporal regions as well as the hippocampus (β=-0.007).

**Figure 3.**
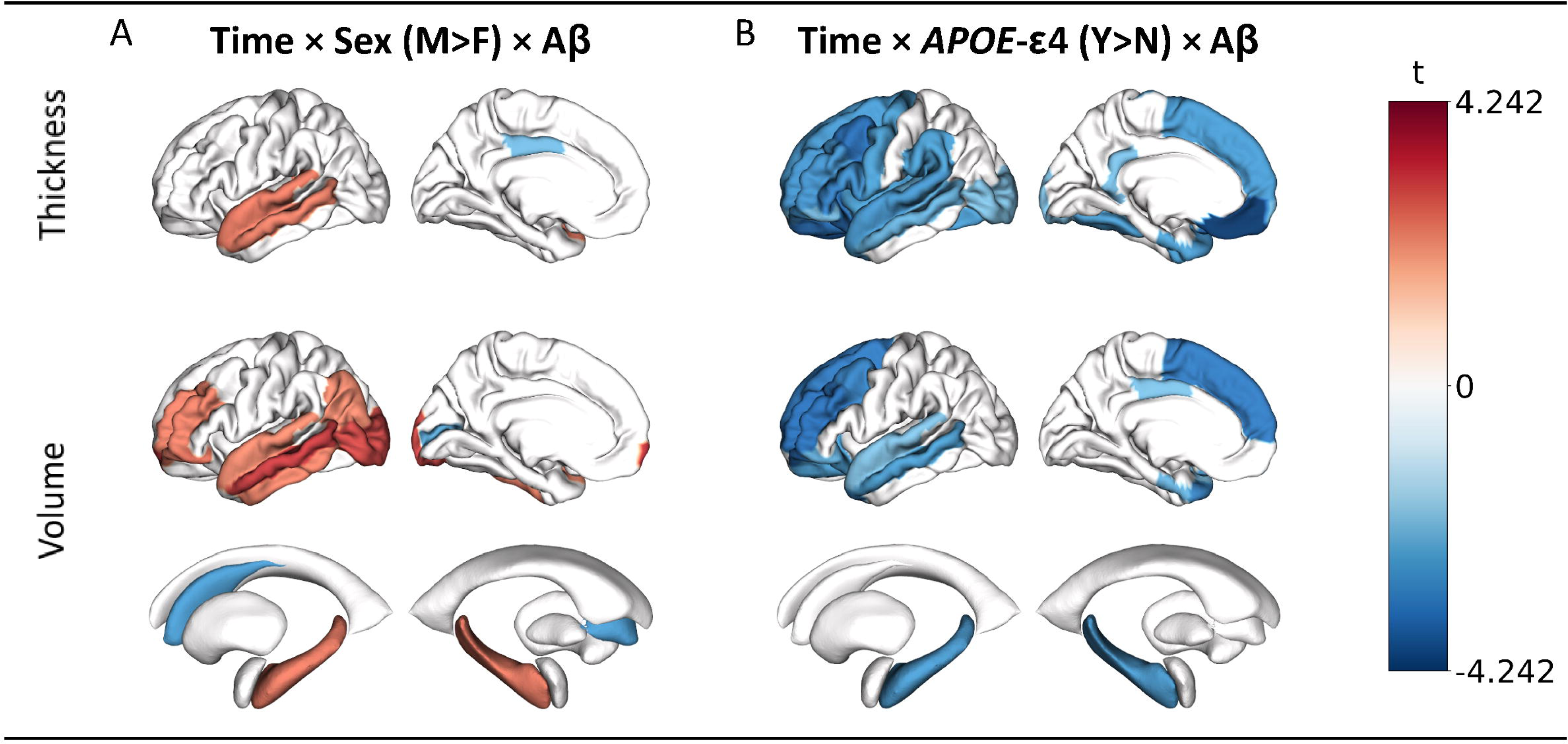
Three-way interaction effects of time, **A**β **burden at baseline and Sex or *APOE*-** ε**4 carriership on regional volumes & thicknesses.** Cortical mid-surface projections of FDR-thresholded LME interaction effect t-values for cortical thickness, cortical volume and subcortical volume, of **(A)** time*Aβ*sex, showing increased lateral and medial temporal atrophy for female compare to male participants with increases in Aβ, and **(B)** time*Aβ\**APOE*-ε4, showing exacerbated atrophy for *APOE*-ε4 carriers in frontal and temporal regions. Surface projections show the lateral and medial cortex respectively, representative of averaged bilateral effects. Aβ, amyloid β.

### 3.3 Follow-Up A**β**-PET data does not improve brain atrophy predictions

LME results of the data subset (*N*=684) with available longitudinal Aβ-PET are shown in **Supplementary** Fig. 5A. Including follow-up PET scans generally did not improve predictions of volumetric or thickness measures, with significant improvements only in the prediction of volume of the posterior cingulate (ΔAIC = −9.55), caudal-anterior cingulate (ΔAIC = −4.97) and superior-frontal cortices (ΔAIC = −2.75; **Supplementary** Fig. 5B).

## 4. Discussion

In this longitudinal prospective pan-European cohort study, we investigated the effect of baseline cortical Aβ burden on subsequent cortical and subcortical brain atrophy in a large cohort of older adults with mostly preserved cognition. We showed that baseline Aβ was predictive of widespread atrophy in several regions, most notably the medial-temporal lobes, fusiform, cingulate, and precuneus cortices, with thickness being a more sensitive or affected measure than volume. When correcting for p-tau or t-tau in a subset with available CSF data, only fusiform volume reductions were predicted by Aβ. Further, we demonstrated that amyloid-induced atrophy is exacerbated in women and with *APOE*-ε4 carriership.

The observed widespread atrophy suggests an early effect of Aβ on brain volume and thickness, and corroborates the importance of early intervention to prevent subsequent atrophy (Aisen et al., 2020). Amyloid-related longitudinal changes were more strongly observed on measures of cortical thickness, and most prominently in the fusiform, precuneus, and superior temporal cortices, matching those ROI’s previously selected in a cortical thickness meta-ROI to distinguish MCI/AD from healthy controls (Schwarz et al., 2016). In contrast, previous whole-brain investigations of the relationship between Aβ and atrophy in preclinical or prodromal AD cohorts have largely focused on volume (Chételat et al., 2012; Tosun et al., 2011). These studies revealed similar affected regions, such as the precuneus, medial and lateral-temporal cortices, and posterior and middle cingulate. Clinical anti-amyloid trials mostly use global, lateral ventricle or hippocampal volumes as secondary outcomes (Rafii et al., 2023), and our results support the use of lateral ventricular and hippocampal volume to be potential surrogate outcomes in secondary prevention trials for subjects with elevated, but not intermediate levels of Aβ burden. Importantly, baseline Aβ remained predictive of fusiform atrophy after correcting for CSF p-tau181 or t-tau concentrations, suggesting a potential route independent of neurofibrillary tangles specifically for the fusiform gyrus. Corroborating these results, a greater atrophy rate in the left fusiform independent of CSF p-tau181 was also previously found when comparing A-T-participants with normal cognition to A+T-participants, with this effect absent in A+T+ individuals compared to either group (Cacciaglia et al., 2025). Conversely, the effect of Aβ on regional atrophy was lost for all other regions when correcting for p-tau181 or t-tau, indicating that even in this early cohort, most of our observed atrophy is probably driven by the presence of fibrillary tau. A caveat of these sensitivity analyses is that p-tau181 is not only representative of neurofibrillary tangles, but also related to Aβ burden itself (Salvadó et al., 2023), whilst t-tau is related to acute neuronal damage (Blennow and Zetterberg, 2018; Skillbäck et al., 2014); hence using these CSF markers as covariates may result in overcorrection. Novel CSF markers such as MTBR-tau243 have been found to be more specifically representative of neurofibrillary tangles and strongly linked to tau-PET burden (Horie et al., 2023). Future work, should investigate the predictive value of Aβ-PET for subsequent atrophy after correcting for MTBR-tau243.

When looking at the interacting effect of common candidate stratification factors in clinical trials, we found widespread sex-dependent effects with stronger amyloid-related atrophy in women, especially in medial and lateral temporal regions. Sauty and Durrleman (2023) previously found near-identical patterns of faster pace of atrophy in women in people with clinical diagnoses of AD, but not in MCI or cognitively unimpaired individuals, suggesting such an interaction effect to be predominant in late disease stages. Our present findings stem from a predominantly cognitively normal population, suggesting that clinical status may not drive accelerated atrophy in women as much as elevated pathological burden of at least Aβ, but likely also neurofibrillary tau: at comparable prevalence rates of tau-PET positivity between men and women (Ossenkoppele et al., 2025), women have been found to have larger quantities of neurofibrillary tangles in temporoparietal regions (Pereira et al., 2020) and faster rate of accumulation in inferior temporal and fusiform regions (Coughlan et al., 2025), suggesting that equal levels of Aβ may lead to more neurofibrillary tau in women, leading subsequently to increased atrophy. We further observed that Aβ-related atrophy is accelerated in *APOE-*ε4 carriers, predominantly in frontal and temporal regions. Such *APOE-*ε4-related atrophy exacerbation has also been found in patients with AD, and in the superior temporal cortex also in people with MCI (Sauty and Durrleman, 2023). *APOE*-ε4 predominance has also previously been reported in an AD frontal subtype of Aβ accumulation (L. E. Collij et al., 2022); which, in light of our findings, could be suggestive of a spatial association between Aβ deposition and downstream atrophy.

In the context of clinical trials, while a reduction of region-specific atrophy, namely hippocampal and lateral ventricular volume loss and fusiform thickness, could theoretically be the most detectable in patients with elevated Aβ levels over 40 CL, existing successful anti-amyloid trials have reported either insignificant or accelerated volume decreases (Alves et al., 2023). These unexpected findings might be caused by off-target reduction of inflammatory responses, resulting in “pseudo-atrophy” (Barkhof and Knopman, 2023), with later discussions observing that such findings majorly occur in otherwise successful trials, designating it more specifically as “amyloid-removal related pseudo-atrophy” (Belder et al., 2024). While our findings highlight the importance of early anti-amyloid treatment to prevent detectable atrophy, atrophy itself would therefore likely not be a good outcome measure in anti-amyloid trials.

Our findings represent the largest population to date to address the relationship between Aβ and atrophy in preclinical AD, enabling the inclusion of relevant covariates and stratification analyses such as the concurrent investigation of regional cortical volume and thickness. However, several methodological limitations need to be considered. First, the lack of a direct marker for neurofibrillary tangles, more closely related to patterns of atrophy in clinical AD than cortical Aβ (Gordon et al., 2018), did not allow us to disentangle the individual contributions of these pathological factors to brain atrophy. However, the observed associations in the CSF p-tau181 and t-tau sensitivity analyses as well as CDR=0 subset analyses with a presumably relatively lower tau burden advocate for a partially independent effect of Aβ on neurodegeneration. The previously observed additive effect of vascular risk to the association between Aβ and atrophy was also not taken into account in this work and warrants future research (Rabin et al., 2022). Finally, the availability of follow-up data was skewed towards low Aβ burden participants, with over 80% of follow-up data coming from the low Aβ group at baseline (**Supplementary** Fig. 2). This bias may explain the overall lack of model improvement when including longitudinal PET data, as previous studies have highlighted the benefits of using follow-up PET data (Jagust et al., 2021). While the acquired data originated from 17 European sites and was acquired with different MRI scanners and Aβ-PET radiotracers, possible batch effects were minimized by utilizing state-of-the-art harmonization techniques (Fortin et al., 2018; Klunk et al., 2015). In the present study, we investigated regional differences through the gyral based Desikan-Killiany atlas; future research could focus on more fine-grained parcellations such as hippocampal subfields, with more focal CA1, presubiculum and subiculum atrophy having been shown to occur before other subfields in the amyloid cascade (Göschel et al., 2023; Zilioli et al., 2025). Additionally, as the global Centiloid metric is a robust measure (Shekari et al., 2024) and commonly used in previous and ongoing trials, we did not investigate regional measures of Aβ burden. However, evidence exists regarding the presence of different amyloid-PET spatial-temporal subtypes (L. E. Collij et al., 2022). It would be of interest to investigate whether regional amyloid-PET quantifications further improve atrophy prediction models. Beyond these aspects, rather than assuming a single pattern of atrophy due to Aβ, subtypes of atrophy (Risacher et al., 2017) in relation to Aβ accumulation could be investigated.

### 4.1 Conclusions

We demonstrated the influence of cortical Aβ on brain atrophy in several regions in a non-demented population. Importantly, this influence was already exerted in cognitively unimpaired individuals and appeared to be mostly independent of sex and *APOE* ε4-genotype. Our results highlight cortical thickness as a more affected biomarker compared to volume. Overall, our findings emphasize the importance of early intervention strategies to mitigate Aβ-related neurodegeneration, with implications for the design and interpretation of clinical trials aimed at preventing AD.

## Data availability

Data for this study were drawn from the AMYPAD PNHS v202306, available on the AD Workbench and can be requested via the AD Workbench FAIR portal under https://fair.addi.addatainitiative.org/#/data/datasets/amypad_pnhsharmonised_and_derivedv202306. The data access request procedure is described under https://doi.org/10.5281/zenodo.7962924.

## Data Availability

Data for this study were drawn from the AMYPAD PNHS v202306, available on the AD Workbench and can be requested via the AD Workbench FAIR portal under https://community.addi.ad-datainitiative.org/datasets/a/d/c/datasets/DA63/amypad-pnhs-harmonised-and-derived-v202306-1865976463. The data access request procedure is described under https://doi.org/10.5281/zenodo.7962924.

https://amypad.eu/data/

## Notes

### Competing Interest Statement

Robin Wolz reports a relationship with IXICO plc that includes: employment. Sylke Grootoonk reports a relationship with IXICO plc that includes: employment. Craig Ritchie reports a relationship with Scottish Brain Sciences that includes: board membership and equity or stocks. Craig Ritchie reports a relationship with Biogen Inc that includes: consulting or advisory fees. Craig Ritchie reports a relationship with Eisai Inc that includes: consulting or advisory fees. Craig Ritchie reports a relationship with MSD that includes: consulting or advisory fees. Craig Ritchie reports a relationship with Actinogen that includes: consulting or advisory fees. Craig Ritchie reports a relationship with Roche that includes: consulting or advisory fees. Craig Ritchie reports a relationship with Eli Lilly and Company that includes: consulting or advisory fees. Merce Boada reports a relationship with Grifols that includes: consulting or advisory fees, board membership, and speaking and lecture fees. Merce Boada reports a relationship with Araclon Biotech that includes: consulting or advisory fees, board membership, and speaking and lecture fees. Merce Boada reports a relationship with Roche that includes: consulting or advisory fees, board membership, and speaking and lecture fees Merce Boada reports a relationship with Biogen that includes: consulting or advisory fees and board membership. Merce Boada reports a relationship with Eli Lilly that includes: consulting or advisory fees and board membership. Merce Boada reports a relationship with Merck that includes: consulting or advisory fees and board membership. Merce Boada reports a relationship with Zambon that includes: consulting or advisory fees and board membership. Merce Boada reports a relationship with Novo Nordisk that includes: consulting or advisory fees, board membership, and speaking and lecture fees. Merce Boada reports a relationship with Bioiberica that includes: board membership. Merce Boada reports a relationship with Eisai that includes: board membership. Merce Boada reports a relationship with Servier that includes: board membership and speaking and lecture fees. Merce Boada reports a relationship with Schwabe Pharma that includes: board membership. Merce Boada reports a relationship with Nutricia that includes: speaking and lecture fees. Marta Marquie reports a relationship with Instituto de Salud Carlos III (ISCIII): funding or grants. Marta Marquie reports a relationship with F. Hoffmann-La Roche Ltd.: travel reimbursement. Marta Marquie reports a relationship with Araclon Biotech-Grifols that includes: board membership. Wiesje van der Flier reports a relationship with Oxford Health Policy Forum CIC: consulting or advisory fees. Wiesje van der Flier reports a relationship with Roche: consulting or advisory fees. Wiesje van der Flier reports a relationship with Biogen MA Inc: consulting or advisory fees. Wiesje van der Flier reports a relationship with Eisai: consulting or advisory fees. Wiesje van der Flier reports a relationship with Biogen MA Inc, Danone, Eisai, WebMD Neurology, Novo Nordisk, and Springer Healthcare that includes: speaking and lecture fees. Wiesje van der Flier reports a relationship with Roche and Eli Lilly that includes: board membership. Wiesje van der Flier reports a relationship with Novo Nordisk that includes: board membership. Wiesje van der Flier reports a relationship with PAVE and Think Brain Health that includes: board membership. Bernard J Hanseeuw reports a relationship with Biogen that includes: consulting or advisory fees. Bernard J Hanseeuw reports a relationship with Eisai that includes: consulting or advisory fees. Bernard J Hanseeuw reports a relationship with Roche that includes: consulting or advisory fees. Pablo Martinez-Lage reports a relationship with Eli Lilly that includes: board membership, speaking and lecture fees, and travel reimbursement. Pablo Martinez-Lage reports a relationship with Nutricia that includes: board membership, speaking and lecture fees, and travel reimbursement. Pablo Martinez-Lage reports a relationship with Roche that includes: board membership, speaking and lecture fees, and travel reimbursement. Pablo Martinez-Lage reports a relationship with Eisai that includes: board membership, speaking and lecture fees, and travel reimbursement. Pablo Martinez-Lage reports a relationship with Grifols that includes: board membership, speaking and lecture fees, and travel reimbursement. Pablo Martinez-Lage reports a relationship with Esteve that includes: board membership, speaking and lecture fees, and travel reimbursement. Pierre Payoux reports a relationship with GE Healthcare that includes: consulting or advisory fees and research support. Pieter Jelle Visser reports a relationship with Eli Lilly that includes: board membership. Pieter Jelle Visser reports a relationship with Janssen Pharmaceuticals that includes: consulting or advisory fees. Pieter Jelle Visser reports a relationship with Bristol-Myers Squibb that includes: Funding or grants. Pieter Jelle Visser reports a relationship with GE Healthcare that includes: Funding or grants. Pieter Jelle Visser reports a relationship with the European Commission that includes: Funding or grants. Pieter Jelle Visser reports a relationship with other European funding programs that includes: Funding or grants. Michael Schoell reports a relationship with Knut and Alice Wallenberg Foundation,the Swedish Research Council, European Union programs that includes: Funding or grants. Michael Schoell reports a relationship with the Swedish Research Council that includes: Funding or grants. Michael Schoell reports a relationship with European Union programs that includes: Funding or grants. Michael Schoell reports a relationship with Roche that includes: board membership. Michael Schoell reports a relationship with Novo Nordisk that includes: board membership. Michael Schoell reports a relationship with Bioarctic that includes: speaking and lecture fees. Michael Schoell reports a relationship with Eisai that includes: speaking and lecture fees. Michael Schoell reports a relationship with Genentech that includes: speaking and lecture fees. Michael Schoell reports a relationship with Novo Nordisk that includes: speaking and lecture fees. Michael Schoell reports a relationship with Roche that includes: speaking and lecture fees. Andrew W. Stephens Collij reports a relationship with Life Molecular Imaging that includes: employment. Christopher Buckley reports a relationship with GE Healthcare that includes: employment. Gill Farrar reports a relationship with GE Healthcare that includes: employment. Lyduine E. Collij reports a relationship with GE Healthcare and Springer Healthcare that includes: research support. If there are other authors, they declare that they have no known competing financial interests or personal relationships that could have appeared to influence the work reported in this paper.

### Clinical Protocols

https://www.clinicaltrialsregister.eu/ctr-search/trial/2018-002277-22/NL

https://onderzoekmetmensen.nl/en/trial/55776

### Funding Statement

The project leading to this paper has received funding from the Innovative Medicines Initiative 2 Joint Undertaking under grant agreement no. 115952. This Joint Undertaking receives the support from the European Union's Horizon 2020 research and innovation programme and EFPIA.
This study was supported by the Alzheimer's Disease Data Initiative (ADDI).

### Author Declarations

The AMYPAD PNHS (EudraCT: 2018-002277-22) is a multi-center cohort of non-demented participants to determine the value of amyloid PET in clinical- and research settings; The individual studies were reviewed and approved by Medical Ethical Committee of the University Medical Center Amsterdam, location VUmc and all local sites.

### Summary of Updates

The term "non-demented" has been eliminated and updated with "older adults without dementia" or similar throughout the manuscript. APOE-e4 and sex interaction analyses have substantially changed due to keeping both interaction terms in the same model rather than separating. Trial power analyses have been removed as these are unlikely to have much scientific value. Results with CDR=0 are now more highlighted, and analyses with CSF p-tau and t-tau corrections have been added to the manuscript.

## References

Aisen, P.S., Cummings, J., Doody, R., Kramer, L., Salloway, S., Selkoe, D.J., Sims, J., Sperling, R.A., Vellas, B., 2020. The Future of Anti-Amyloid Trials. J Prev Alzheimers Dis 7, 146–151. 10.14283/jpad.2020.24

Alves, F., Kalinowski, P., Ayton, S., 2023. Accelerated Brain Volume Loss Caused by Anti-β-Amyloid Drugs: A Systematic Review and Meta-analysis. Neurology 100, e2114– e2124. 10.1212/WNL.0000000000207156

Barkhof, F., Knopman, D.S., 2023. Brain Shrinkage in Anti–β-Amyloid Alzheimer Trials: Neurodegeneration or Pseudoatrophy?: Neurology: Vol 100, No 20. Neurology 100, 941–942. 10.1212/WNL.0000000000207268

Barthel, H., Gertz, H.-J., Dresel, S., Peters, O., Bartenstein, P., Buerger, K., Hiemeyer, F., Wittemer-Rump, S.M., Seibyl, J., Reininger, C., Sabri, O., Florbetaben Study Group, 2011. Cerebral amyloid-β PET with florbetaben (18F) in patients with Alzheimer’s disease and healthy controls: a multicentre phase 2 diagnostic study. Lancet Neurol. 10, 424–435. 10.1016/S1474-4422(11)70077-1

Bejanin, A., Schonhaut, D.R., La Joie, R., Kramer, J.H., Baker, S.L., Sosa, N., Ayakta, N., Cantwell, A., Janabi, M., Lauriola, M., O’Neil, J.P., Gorno-Tempini, M.L., Miller, Z.A., Rosen, H.J., Miller, B.L., Jagust, W.J., Rabinovici, G.D., 2017. Tau pathology and neurodegeneration contribute to cognitive impairment in Alzheimer’s disease. Brain 140, 3286–3300. 10.1093/brain/awx243

Belder, C.R.S., Boche, D., Nicoll, J.A.R., Jaunmuktane, Z., Zetterberg, H., Schott, J.M., Barkhof, F., Fox, N.C., 2024. Brain volume change following anti-amyloid β immunotherapy for Alzheimer’s disease: amyloid-removal-related pseudo-atrophy. Lancet Neurol. 23, 1025–1034. 10.1016/S1474-4422(24)00335-1

Benjamini, Y., Hochberg, Y., 1995. Controlling the false discovery rate: A practical and powerful approach to multiple testing. J. R. Stat. Soc. 57, 289–300. 10.1111/j.2517-6161.1995.tb02031.x

Blennow, K., Zetterberg, H., 2018. Biomarkers for Alzheimer’s disease: current status and prospects for the future. J. Intern. Med. 284, 643–663. 10.1111/joim.12816

Buckley, R.F., Mormino, E.C., Rabin, J.S., Hohman, T.J., Landau, S., Hanseeuw, B.J., Jacobs, H.I.L., Papp, K.V., Amariglio, R.E., Properzi, M.J., Schultz, A.P., Kirn, D., Scott, M.R., Hedden, T., Farrell, M., Price, J., Chhatwal, J., Rentz, D.M., Villemagne, V.L., Johnson, K.A., Sperling, R.A., 2019. Sex Differences in the Association of Global Amyloid and Regional Tau Deposition Measured by Positron Emission Tomography in Clinically Normal Older Adults. JAMA Neurol. 76, 542–551. 10.1001/jamaneurol.2018.4693

Cacciaglia, R., Falcón, C., Benavides, G.S., Brugulat-Serrat, A., Alomà, M.M., Calvet, M.S., Molinuevo, J.L., Fauria, K., Minguillón, C., Kollmorgen, G., Quijano-Rubio, C., Blennow, K., Zetterberg, H., Lorenzini, L., Wink, A.M., Ingala, S., Barkhof, F., Ritchie, C.W., Gispert, J.D., ALFA study, 2025. Soluble Aβ pathology predicts neurodegeneration and cognitive decline independently on p-tau in the earliest Alzheimer’s continuum: Evidence across two independent cohorts. Alzheimers. Dement. 21, e14415. 10.1002/alz.14415

Chételat, G., Villemagne, V.L., Pike, K.E., Baron, J.-C., Bourgeat, P., Jones, G., Faux, N.G., Ellis, K.A., Salvado, O., Szoeke, C., Martins, R.N., Ames, D., Masters, C.L., Rowe, C.C., Australian Imaging Biomarkers and Lifestyle Study of Ageing (AIBL) Research Group, 2010. Larger temporal volume in elderly with high versus low beta-amyloid deposition. Brain 133, 3349–3358. 10.1093/brain/awq187

Chételat, G., Villemagne, V.L., Villain, N., Jones, G., Ellis, K.A., Ames, D., Martins, R.N., Masters, C.L., Rowe, C.C., AIBL Research Group, 2012. Accelerated cortical atrophy in cognitively normal elderly with high β-amyloid deposition. Neurology 78, 477–484. 10.1212/WNL.0b013e318246d67a

Collij, Lyduine E., Farrar, G., Valléz García, D., Bader, I., Shekari, M., Lorenzini, L., Pemberton, H., Altomare, D., Pla, S., Loor, M., Markiewicz, P., Yaqub, M., Buckley, C., Frisoni, G.B., Nordberg, A., Payoux, P., Stephens, A., Gismondi, R., Visser, P.J., Ford, L., Schmidt, M., Birck, C., Georges, J., Mett, A., Walker, Z., Boada, M., Drzezga, A., Vandenberghe, R., Hanseeuw, B., Jessen, F., Schöll, M., Ritchie, C., Lopes Alves, I., Gispert, J.D., Barkhof, F., 2022. The amyloid imaging for the prevention of Alzheimer’s disease consortium: A European collaboration with global impact. Front. Neurol. 13, 1063598. 10.3389/fneur.2022.1063598

Collij, L.E., Salvadó, G., Shekari, M., Lopes Alves, I., Reimand, J., Wink, A.M., Zwan, M., Niñerola-Baizán, A., Perissinotti, A., Scheltens, P., Ikonomovic, M.D., Smith, A.P.L., Farrar, G., Molinuevo, J.L., Barkhof, F., Buckley, C.J., van Berckel, B.N.M., Gispert, J.D., ALFA study, AMYPAD consortium, 2021. Visual assessment of [18F]flutemetamol PET images can detect early amyloid pathology and grade its extent. Eur. J. Nucl. Med. Mol. Imaging 48, 2169–2182. 10.1007/s00259-020-05174-2

Collij, L. E., Salvadó, G., Wottschel, V., Mastenbroek, S.E., Schoenmakers, P., Heeman, F., Aksman, L., Wink, A.M., Berckel, B.N.M., Flier, W.M., Scheltens, P., Visser, P.J., Barkhof, F., Haller, S., Gispert, J.D., Lopes Alves, I., 2022. Spatial-Temporal Patterns of β-Amyloid Accumulation: A Subtype and Stage Inference Model Analysis. Neurology 98. https://www.neurology.org/doi/10.1212/WNL.0000000000200148

Coughlan, G.T., Klinger, H.M., Boyle, R., Betthauser, T.J., Binette, A.P., Christenson, L., Chadwick, T., Hansson, O., Harrison, T.M., Healy, B., Jacobs, H.I.L., Hanseeuw, B., Jonaitis, E., Jack, C.R., Jr, Johnson, K.A., Langhough, R.E., Properzi, M.J., Rentz, D.M., Schultz, A.P., Smith, R., Seto, M., Johnson, S.C., Mielke, M.M., Shirzadi, Z., Yau, W.-Y.W., Manson, J.E., Sperling, R.A., Vemuri, P., Buckley, R.F., Alzheimer’s Disease Neuroimaging Initiative, 2025. Sex differences in longitudinal tau-PET in preclinical Alzheimer disease: A meta-analysis. JAMA Neurol. 82, 364. 10.1001/jamaneurol.2025.0013

Curtis, C., Gamez, J.E., Singh, U., Sadowsky, C.H., Villena, T., Sabbagh, M.N., Beach, T.G., Duara, R., Fleisher, A.S., Frey, K.A., Walker, Z., Hunjan, A., Holmes, C., Escovar, Y.M., Vera, C.X., Agronin, M.E., Ross, J., Bozoki, A., Akinola, M., Shi, J., Vandenberghe, R., Ikonomovic, M.D., Sherwin, P.F., Grachev, I.D., Farrar, G., Smith, A.P.L., Buckley, C.J., McLain, R., Salloway, S., 2015. Phase 3 trial of flutemetamol labeled with radioactive fluorine 18 imaging and neuritic plaque density. JAMA Neurol. 72, 287–294. 10.1001/jamaneurol.2014.4144

Desikan, R.S., Ségonne, F., Fischl, B., Quinn, B.T., Dickerson, B.C., Blacker, D., Buckner, R.L., Dale, A.M., Maguire, R.P., Hyman, B.T., Albert, M.S., Killiany, R.J., 2006. An automated labeling system for subdividing the human cerebral cortex on MRI scans into gyral based regions of interest. Neuroimage 31, 968–980. 10.1016/j.neuroimage.2006.01.021

Dhamala, E., Ooi, L.Q.R., Chen, J., Kong, R., Anderson, K.M., Chin, R., Yeo, B.T.T., Holmes, A.J., 2022. Proportional intracranial volume correction differentially biases behavioral predictions across neuroanatomical features, sexes, and development. Neuroimage 260, 119485. 10.1016/j.neuroimage.2022.119485

Fortin, J.-P., Cullen, N., Sheline, Y.I., Taylor, W.D., Aselcioglu, I., Cook, P.A., Adams, P., Cooper, C., Fava, M., McGrath, P.J., McInnis, M., Phillips, M.L., Trivedi, M.H., Weissman, M.M., Shinohara, R.T., 2018. Harmonization of cortical thickness measurements across scanners and sites. Neuroimage 167, 104–120. 10.1016/j.neuroimage.2017.11.024

García, D.V., 2023. AMYPAD PNHS - Integrated dataset (Raw) v202306. 10.5281/ZENODO.8017084

Gordon, B.A., McCullough, A., Mishra, S., Blazey, T.M., Su, Y., Christensen, J., Dincer, A., Jackson, K., Hornbeck, R.C., Morris, J.C., Ances, B.M., Benzinger, T.L.S., 2018. Cross-sectional and longitudinal atrophy is preferentially associated with tau rather than amyloid β positron emission tomography pathology. Alzheimers. Dement. 10, 245–252. 10.1016/j.dadm.2018.02.003

Göschel, L., Kurz, L., Dell’Orco, A., Köbe, T., Körtvélyessy, P., Fillmer, A., Aydin, S., Riemann, L.T., Wang, H., Ittermann, B., Grittner, U., Flöel, A., 2023. 7T amygdala and hippocampus subfields in volumetry-based associations with memory: A 3-year follow-up study of early Alzheimer’s disease. NeuroImage Clin. 38, 103439. 10.1016/j.nicl.2023.103439

Gustavsson, A., Norton, N., Fast, T., Frölich, L., Georges, J., Holzapfel, D., Kirabali, T., Krolak-Salmon, P., Rossini, P.M., Ferretti, M.T., Lanman, L., Chadha, A.S., van der Flier, W.M., 2023. Global estimates on the number of persons across the Alzheimer’s disease continuum. Alzheimers. Dement. 19, 658–670. 10.1002/alz.12694

Horie, K., Salvadó, G., Barthélemy, N.R., Janelidze, S., Li, Y., He, Y., Saef, B., Chen, C.D., Jiang, H., Strandberg, O., Pichet Binette, A., Palmqvist, S., Sato, C., Sachdev, P., Koyama, A., Gordon, B.A., Benzinger, T.L.S., Holtzman, D.M., Morris, J.C., Mattsson-Carlgren, N., Stomrud, E., Ossenkoppele, R., Schindler, S.E., Hansson, O., Bateman, R.J., 2023. CSF MTBR-tau243 is a specific biomarker of tau tangle pathology in Alzheimer’s disease. Nat. Med. 29, 1954–1963. 10.1038/s41591-023-02443-z

Ingala, S., De Boer, C., Masselink, L.A., Vergari, I., Lorenzini, L., Blennow, K., Chételat, G., Di Perri, C., Ewers, M., van der Flier, W.M., Fox, N.C., Gispert, J.D., Haller, S., Molinuevo, J.L., Muniz-Terrera, G., Mutsaerts, H.J., Ritchie, C.W., Ritchie, K., Schmidt, M., Schwarz, A.J., Vermunt, L., Waldman, A.D., Wardlaw, J., Wink, A.M., Wolz, R., Wottschel, V., Scheltens, P., Visser, P.J., Barkhof, F., EPAD consortium, 2021. Application of the ATN classification scheme in a population without dementia: Findings from the EPAD cohort. Alzheimers. Dement. 17, 1189–1204. 10.1002/alz.12292

Jack, C.R., Jr, Wiste, H.J., Weigand, S.D., Therneau, T.M., Lowe, V.J., Knopman, D.S., Gunter, J.L., Senjem, M.L., Jones, D.T., Kantarci, K., Machulda, M.M., Mielke, M.M., Roberts, R.O., Vemuri, P., Reyes, D.A., Petersen, R.C., 2017. Defining imaging biomarker cut points for brain aging and Alzheimer’s disease. Alzheimers. Dement. 13, 205–216. 10.1016/j.jalz.2016.08.005

Jagust, W.J., Landau, S.M., Alzheimer’s Disease Neuroimaging Initiative, 2021. Temporal Dynamics of β-Amyloid Accumulation in Aging and Alzheimer Disease. Neurology 96, e1347–e1357. 10.1212/WNL.0000000000011524

Josephs, K.A., Whitwell, J.L., Ahmed, Z., Shiung, M.M., Weigand, S.D., Knopman, D.S., Boeve, B.F., Parisi, J.E., Petersen, R.C., Dickson, D.W., Jack, C.R., Jr, 2008. Beta-amyloid burden is not associated with rates of brain atrophy. Ann. Neurol. 63, 204–212. 10.1002/ana.21223

Kepp, K.P., Sensi, S.L., Johnsen, K.B., Barrio, J.R., Høilund-Carlsen, P.F., Neve, R.L., Alavi, A., Herrup, K., Perry, G., Robakis, N.K., Vissel, B., Espay, A.J., 2023. The Anti-Amyloid Monoclonal Antibody Lecanemab: 16 Cautionary Notes. J. Alzheimers. Dis. 94, 497–507. 10.3233/JAD-230099

Klunk, W.E., Koeppe, R.A., Price, J.C., Benzinger, T.L., Devous, M.D., Sr, Jagust, W.J., Johnson, K.A., Mathis, C.A., Minhas, D., Pontecorvo, M.J., Rowe, C.C., Skovronsky, D.M., Mintun, M.A., 2015. The Centiloid Project: standardizing quantitative amyloid plaque estimation by PET. Alzheimers. Dement. 11, 1–15.e1–4. 10.1016/j.jalz.2014.07.003

La Joie, R., Visani, A.V., Baker, S.L., Brown, J.A., Bourakova, V., Cha, J., Chaudhary, K., Edwards, L., Iaccarino, L., Janabi, M., Lesman-Segev, O.H., Miller, Z.A., Perry, D.C., O’Neil, J.P., Pham, J., Rojas, J.C., Rosen, H.J., Seeley, W.W., Tsai, R.M., Miller, B.L., Jagust, W.J., Rabinovici, G.D., 2020. Prospective longitudinal atrophy in Alzheimer’s disease correlates with the intensity and topography of baseline tau-PET. Sci. Transl. Med. 12. 10.1126/scitranslmed.aau5732

Lopes Alves, I., Collij, L.E., Altomare, D., Frisoni, G.B., Saint-Aubert, L., Payoux, P., Kivipelto, M., Jessen, F., Drzezga, A., Leeuwis, A., Wink, A.M., Visser, P.J., van Berckel, B.N.M., Scheltens, P., Gray, K.R., Wolz, R., Stephens, A., Gismondi, R., Buckely, C., Gispert, J.D., Schmidt, M., Ford, L., Ritchie, C., Farrar, G., Barkhof, F., Molinuevo, J.L., AMYPAD Consortium, 2020. Quantitative amyloid PET in Alzheimer’s disease: the AMYPAD prognostic and natural history study. Alzheimers. Dement. 16, 750–758. 10.1002/alz.12069

Martikainen, I.K., Kemppainen, N., Johansson, J., Teuho, J., Helin, S., Liu, Y., Helisalmi, S., Soininen, H., Parkkola, R., Ngandu, T., Kivipelto, M., Rinne, J.O., 2019. Brain β-Amyloid and Atrophy in Individuals at Increased Risk of Cognitive Decline. AJNR Am. J. Neuroradiol. 40, 80–85. 10.3174/ajnr.A5891

Mattsson, N., Eriksson, O., Lindberg, O., Schöll, M., Lampinen, B., Nilsson, M., Insel, P.S., Lautner, R., Strandberg, O., van Westen, D., Zetterberg, H., Blennow, K., Palmqvist, S., Stomrud, E., Hansson, O., 2018. Effects of APOE ε4 on neuroimaging, cerebrospinal fluid biomarkers, and cognition in prodromal Alzheimer’s disease. Neurobiol. Aging 71, 81–90. 10.1016/j.neurobiolaging.2018.07.003

Mattsson, N., Insel, P.S., Nosheny, R., Tosun, D., Trojanowski, J.Q., Shaw, L.M., Jack, C.R., Jr, Donohue, M.C., Weiner, M.W., Alzheimer’s Disease Neuroimaging Initiative, 2014. Emerging β-amyloid pathology and accelerated cortical atrophy. JAMA Neurol. 71, 725–734. 10.1001/jamaneurol.2014.446

Ossenkoppele, R., Coomans, E.M., Apostolova, L.G., Baker, S.L., Barthel, H., Beach, T.G., Benzinger, T.L.S., Betthauser, T., Bischof, G.N., Bottlaender, M., Bourgeat, P., den Braber, A., Brendel, M., Brickman, A.M., Cash, D.M., Carrillo, M.C., Coath, W., Christian, B.T., Dickerson, B.C., Dore, V., Drzezga, A., Feizpour, A., van der Flier, W.M., Franzmeier, N., Frisoni, G.B., Garibotto, V., van de Giessen, E., Domingo-Gispert, J., Gnoerich, J., Gu, Y., Guan, Y., Hanseeuw, B.J., Harrison, T.M., Jack, C.R., Jaeger, E., Jagust, W.J., Jansen, W.J., La Joie, R., Johnson, K.A., Johnson, S.C., Kennedy, I.A., Kim, J.P., van Laere, K., Lagarde, J., Lao, P., Luchsinger, J.A., Kern, S., Kreisl, W.C., Malotaux, V., Malpetti, M., Manly, J.J., Mao, X., Mattsson-Carlgren, N., Mayo Clinic Study on Aging, Messerschmidt, K., Minguillon, C., Mormino, E.M., O’Brien, J.T., Palmqvist, S., Peretti, D.E., Petersen, R.C., Pijnenburg, Y.A.L., Pontecorvo, M.J., Poirier, J., PREVENT-AD Research Group, Rabinovici, G.D., Rahmouni, N., Risacher, S.L., Rosa-Neto, P., Rosen, H., Rowe, C.C., Rowe, J.B., Rullmann, M., Salman, Y., Sarazin, M., Saykin, A.J., Schneider, J.A., Schöll, M., Schott, J.M., Seo, S.W., Serrano, G.E., Shcherbinin, S., Shekari, M., Skoog, I., Smith, R., Sperling, R.A., Spruyt, L., Stomrud, E., Strandberg, O., Therriault, J., Xie, F., Vandenberghe, R., Villemagne, V.L., Villeneuve, S., Visser, P.J., Vossler, H., Young, C.B., Groot, C., Hansson, O., 2025. Tau PET positivity in individuals with and without cognitive impairment varies with age, amyloid-β status, APOE genotype and sex. Nat. Neurosci. 28, 1610–1621. 10.1038/s41593-025-02000-6

Pereira, J.B., Harrison, T.M., La Joie, R., Baker, S.L., Jagust, W.J., 2020. Spatial patterns of tau deposition are associated with amyloid, ApoE, sex, and cognitive decline in older adults. Eur. J. Nucl. Med. Mol. Imaging 47, 2155–2164. 10.1007/s00259-019-04669-x

Rabin, J.S., Pruzin, J., Scott, M., Yang, H.-S., Hampton, O., Hsieh, S., Schultz, A.P., Buckley, R.F., Hedden, T., Rentz, D., Johnson, K.A., Sperling, R.A., Chhatwal, J.P., 2022. Association of β-amyloid and vascular risk on longitudinal patterns of brain atrophy. Neurology 99, e270–e280. 10.1212/WNL.0000000000200551

Rafii, M.S., Sperling, R.A., Donohue, M.C., Zhou, J., Roberts, C., Irizarry, M.C., Dhadda, S., Sethuraman, G., Kramer, L.D., Swanson, C.J., Li, D., Krause, S., Rissman, R.A., Walter, S., Raman, R., Johnson, K.A., Aisen, P.S., 2023. The AHEAD 3-45 Study: Design of a prevention trial for Alzheimer’s disease. Alzheimers. Dement. 19, 1227– 1233. 10.1002/alz.12748

Reuter, M., Schmansky, N.J., Rosas, H.D., Fischl, B., 2012. Within-subject template estimation for unbiased longitudinal image analysis. Neuroimage 61, 1402–1418. 10.1016/j.neuroimage.2012.02.084

Risacher, S.L., Anderson, W.H., Charil, A., Castelluccio, P.F., Shcherbinin, S., Saykin, A.J., Schwarz, A.J., Alzheimer’s Disease Neuroimaging Initiative, 2017. Alzheimer disease brain atrophy subtypes are associated with cognition and rate of decline. Neurology 89, 2176–2186. 10.1212/WNL.0000000000004670

Salvadó, G., Molinuevo, J.L., Brugulat-Serrat, A., Falcon, C., Grau-Rivera, O., Suárez-Calvet, M., Pavia, J., Niñerola-Baizán, A., Perissinotti, A., Lomeña, F., Minguillon, C., Fauria, K., Zetterberg, H., Blennow, K., Gispert, J.D., Alzheimer’s Disease Neuroimaging Initiative, for the ALFA Study, 2019. Centiloid cut-off values for optimal agreement between PET and CSF core AD biomarkers. Alzheimers. Res. Ther. 11, 27. 10.1186/s13195-019-0478-z

Salvadó, G., Ossenkoppele, R., Ashton, N.J., Beach, T.G., Serrano, G.E., Reiman, E.M., Zetterberg, H., Mattsson-Carlgren, N., Janelidze, S., Blennow, K., Hansson, O., 2023. Specific associations between plasma biomarkers and postmortem amyloid plaque and tau tangle loads. EMBO Mol. Med. 15, e17123. 10.15252/emmm.202217123

Sauty, B., Durrleman, S., 2023. Impact of sex and APOE-ε4 genotype on patterns of regional brain atrophy in Alzheimer’s disease and healthy aging. Front. Neurol. 14, 1161527. 10.3389/fneur.2023.1161527

Schwarz, A.J., Sundell, K.L., Charil, A., Case, M.G., Jaeger, R.K., Scott, D., Bracoud, L., Oh, J., Suhy, J., Pontecorvo, M.J., Dickerson, B.C., Siemers, E.R., 2019. Magnetic resonance imaging measures of brain atrophy from the EXPEDITION3 trial in mild Alzheimer’s disease. Alzheimers. Dement. 5, 328–337. 10.1016/j.trci.2019.05.007

Schwarz, C.G., Gunter, J.L., Wiste, H.J., Przybelski, S.A., Weigand, S.D., Ward, C.P., Senjem, M.L., Vemuri, P., Murray, M.E., Dickson, D.W., Parisi, J.E., Kantarci, K., Weiner, M.W., Petersen, R.C., Jack, C.R., Jr, Alzheimer’s Disease Neuroimaging Initiative, 2016. A large-scale comparison of cortical thickness and volume methods for measuring Alzheimer’s disease severity. NeuroImage Clin. 11, 802–812. 10.1016/j.nicl.2016.05.017

Shekari, M., Vállez García, D., Collij, L.E., Altomare, D., Heeman, F., Pemberton, H., Roé Vellvé, N., Bullich, S., Buckley, C., Stephens, A., Farrar, G., Frisoni, G., Klunk, W.E., Barkhof, F., Gispert, J.D., ADNI and the AMYPAD consortium, 2024. Stress testing the Centiloid: Precision and variability of PET quantification of amyloid pathology. Alzheimers. Dement. 10.1002/alz.13883

Skillbäck, T., Rosén, C., Asztely, F., Mattsson, N., Blennow, K., Zetterberg, H., 2014. Diagnostic performance of cerebrospinal fluid total tau and phosphorylated tau in Creutzfeldt-Jakob disease: results from the Swedish Mortality Registry. JAMA Neurol. 71, 476–483. 10.1001/jamaneurol.2013.6455

Tosun, D., Schuff, N., Mathis, C.A., Jagust, W., Weiner, M.W., Alzheimer’s Disease NeuroImaging Initiative, 2011. Spatial patterns of brain amyloid-beta burden and atrophy rate associations in mild cognitive impairment. Brain 134, 1077–1088. 10.1093/brain/awr044

Villeneuve, S., Rabinovici, G.D., Cohn-Sheehy, B.I., Madison, C., Ayakta, N., Ghosh, P.M., La Joie, R., Arthur-Bentil, S.K., Vogel, J.W., Marks, S.M., Lehmann, M., Rosen, H.J., Reed, B., Olichney, J., Boxer, A.L., Miller, B.L., Borys, E., Jin, L.-W., Huang, E.J., Grinberg, L.T., DeCarli, C., Seeley, W.W., Jagust, W., 2015. Existing Pittsburgh Compound-B positron emission tomography thresholds are too high: statistical and pathological evaluation. Brain 138, 2020–2033. 10.1093/brain/awv112

Wisch, J.K., Meeker, K.L., Gordon, B.A., Flores, S., Dincer, A., Grant, E.A., Benzinger, T.L., Morris, J.C., Ances, B.M., 2021. Sex-related Differences in Tau Positron Emission Tomography (PET) and the Effects of Hormone Therapy (HT). Alzheimer Dis. Assoc. Disord. 35, 164–168. 10.1097/WAD.0000000000000393

Wolz, R., Aljabar, P., Hajnal, J.V., Hammers, A., Rueckert, D., Alzheimer’s Disease Neuroimaging Initiative, 2010. LEAP: learning embeddings for atlas propagation. Neuroimage 49, 1316–1325. 10.1016/j.neuroimage.2009.09.069

Zilioli, A., Pancaldi, B., Baumeister, H., Busi, G., Misirocchi, F., Mutti, C., Florindo, I., Morelli, N., Mohanty, R., Berron, D., Westman, E., Spallazzi, M., 2025. Unveiling the hippocampal subfield changes across the Alzheimer’s disease continuum: a systematic review of neuroimaging studies. Brain Imaging Behav. 19, 253–267. 10.1007/s11682-024-00952-0

