## Supplementary Material for "Amyloid PET predicts atrophy in older adults without dementia: Results from the AMYPAD Prognostic & Natural History Study"

**Supplement Content**

**Table 1**. Demographics and Clinical Characteristics Stratified by Sex

**Table 2.** Demographics and Clinical Characteristics Stratified by APOE ε4-Carriership

**Table 3.** Regional effects of time and amyloid on volume/thickness

**Table 4.** Effects of time and amyloid on volume/thickness in participants with CSF data

**Figure 1.** Hippocampal volume before and after harmonization with NeuroComBat

**Figure 2.** Distribution of Participants in Amyloid Groups

**Figure 3.** Main results replicated across amyloid stages

**Figure 4.** Longitudinal-only models and model fit improvement with time-varying Aβ

| **Table 1**. **Demographics and Clinical Characteristics Stratified by Sex** | | | | |
| --- | --- | --- | --- | --- |
| **Variable** | **Overall**, N = 1,329^a^ | **Female**, N = 741^a^ | **Male**, N = 588^a^ | **p-value** |
| Age, years | 68.18 (8.78) | 67.68 (9.20) | 68.81 (8.19) | 0.001^b^ |
| Follow-Up, years | 3.74 (1.87) | 3.74 (1.79) | 3.73 (1.97) | 0.5^b^ |
| (Missing) | 585 | 317 | 268 |  |
| Education, years | 14.56 (3.97) | 13.80 (3.86) | 15.53 (3.90) | <0.001^b^ |
| (Missing) | 3 | 2 | 1 |  |
| CDR: 0 - Normal | 1,070 (81%) | 622 (84%) | 448 (76%) | <0.001^c^ |
| MMSE | 28.82 (1.54) | 28.86 (1.51) | 28.76 (1.57) | 0.2^b^ |
| (Missing) | 108 | 60 | 48 |  |
| APOE ε4 carriership (% carriers) | 517 (40%) | 283 (39%) | 234 (41%) | 0.4^c^ |
| (Missing) | 40 | 18 | 22 |  |
| Amyloid PET, centiloids | 19.51 (32.33) | 18.38 (31.47) | 20.94 (33.36) | 0.2^b^ |
| Amyloid Stage |  |  |  | 0.2^c^ |
| Low Aβ | 952 (72%) | 545 (74%) | 407 (69%) |  |
| Intermediate Aβ | 117 (9%) | 63 (9%) | 54 (9%) |  |
| Elevated Aβ | 260 (20%) | 133 (18%) | 127 (22%) |  |
| est. Total Intracranial Volume, mm³ | 1477.71 (178.43) | 1390.51 (146.88) | 1587.60 (152.10) | <0.001^b^ |
| ^a^Mean (SD); n (%) | | | | |
| ^b^Wilcoxon rank sum test | | | | |
| ^c^Pearson's Chi-squared test | | | | |
| Abbreviations: Aβ, amyloid β; CDR, Clinical Dementia Rating; MMSE, Mini-Mental State Examination; PET, Positron Emission Tomography; Amyloid stages are defined based on a Centiloid value of under 20 for low, over 40 for elevated Aβ and a subsequent intermediate Aβ between 20 and 40 CL | | | | |

| **Table 2.** **Demographics and Clinical Characteristics Stratified by APOE ε4-Carriership** | | | | |
| --- | --- | --- | --- | --- |
| **Variable** | **Overall**, N = 1,289^a^ | **No**, N = 772^a^ | **Yes**, N = 517^a^ | **p-value** |
| Age, years | 68.03 (8.71) | 69.02 (9.12) | 66.56 (7.85) | <0.001^b^ |
| Follow-Up, years | 3.77 (1.87) | 3.85 (1.95) | 3.66 (1.74) | 0.2^b^ |
| (Missing) | 563 | 351 | 212 |  |
| Gender: Female (%) | 723 (56%) | 440 (57%) | 283 (55%) | 0.4^c^ |
| Education, years | 14.60 (3.97) | 14.66 (4.04) | 14.52 (3.86) | 0.7^b^ |
| (Missing) | 3 | 1 | 2 |  |
| CDR: 0 - Normal | 1,051 (82%) | 651 (84%) | 400 (77%) | 0.002^c^ |
| MMSE | 28.86 (1.45) | 28.99 (1.23) | 28.67 (1.71) | 0.019^b^ |
| (Missing) | 98 | 64 | 34 |  |
| Amyloid PET, centiloids | 18.65 (31.22) | 11.33 (24.45) | 29.57 (36.61) | <0.001^b^ |
| Amyloid Stage |  |  |  | <0.001^c^ |
| Low Aβ | 935 (73%) | 635 (82%) | 300 (58%) |  |
| Intermediate Aβ | 114 (9%) | 56 (7%) | 58 (11%) |  |
| Elevated Aβ | 240 (19%) | 81 (10%) | 159 (31%) |  |
| est. Total Intracranial Volume, mm³ | 1476.60 (176.72) | 1481.39 (179.05) | 1469.46 (173.10) | 0.3^b^ |
| ^a^Mean (SD); n (%) | | | | |
| ^b^Wilcoxon rank sum test | | | | |
| ^c^Pearson's Chi-squared test | | | | |
| Abbreviations: Aβ, amyloid β; CDR, Clinical Dementia Rating; MMSE, Mini-Mental State Examination; PET, Positron Emission Tomography; Amyloid stages are defined based on a Centiloid value of under 20 for low, over 40 for elevated Aβ and a subsequent intermediate Aβ between 20 and 40 CL | | | | |

| **Table 3.** **Regional effects of time and amyloid on volume/thickness** | | | | | | |
| --- | --- | --- | --- | --- | --- | --- |
|  | Volume | | | Thickness | | |
|  | Time | Aβ | Time × Aβ | Time | Aβ | Time × Aβ |
| **Frontal** |  |  |  |  |  |  |
| Superior Frontal | **-0.02 (0.004)** | -0.001 (0.006) | -0.001 (0.001) | -0.006 (0.006) | **-0.025 (0.007)** | -0.003 (0.002) |
| Rostral Middle Frontal | **-0.032 (0.003)** | -0.007 (0.006) | 0.001 (0.001) | -0.01 (0.006) | **-0.016 (0.007)** | 0.001 (0.002) |
| Caudal Middle Frontal | **-0.015 (0.003)** | -0.014 (0.007) | 0.001 (0.001) | -0.006 (0.005) | **-0.016 (0.007)** | -0.001 (0.002) |
| Pars Opercularis | **-0.03 (0.003)** | -0.01 (0.007) | -0.001 (0.001) | **-0.028 (0.005)** | **-0.021 (0.007)** | -0.001 (0.002) |
| Pars Orbitalis | **-0.033 (0.004)** | **-0.024 (0.007)** | -0.001 (0.001) | -0.006 (0.006) | **-0.026 (0.007)** | 0 (0.002) |
| Pars Triangularis | **-0.032 (0.003)** | -0.006 (0.007) | -0.001 (0.001) | **-0.019 (0.005)** | **-0.015 (0.007)** | 0 (0.002) |
| Medial Orbitofrontal | **-0.03 (0.004)** | **-0.014 (0.006)** | -0.003 (0.002) | -0.007 (0.007) | **-0.028 (0.007)** | **-0.006 (0.002)** |
| Lateral Orbitofrontal | **-0.041 (0.004)** | **-0.018 (0.006)** | -0.002 (0.001) | **-0.021 (0.006)** | **-0.028 (0.007)** | -0.001 (0.002) |
| Precentral | **-0.018 (0.004)** | 0.004 (0.007) | -0.003 (0.001) | **-0.019 (0.006)** | -0.007 (0.007) | -0.004 (0.002) |
| Paracentral | -0.006 (0.004) | 0.005 (0.007) | -0.003 (0.002) | -0.001 (0.006) | -0.006 (0.008) | -0.004 (0.002) |
| Frontal Pole | -0.011 (0.007) | -0.003 (0.007) | 0.003 (0.002) | -0.013 (0.007) | **-0.018 (0.007)** | 0.002 (0.002) |
| Rostral Anterior Cingulate | **-0.021 (0.003)** | -0.015 (0.007) | 0 (0.001) | **0.028 (0.006)** | -0.016 (0.008) | -0.004 (0.002) |
| Caudal Anterior Cingulate | **-0.013 (0.003)** | 0 (0.008) | **-0.003 (0.001)** | **0.021 (0.005)** | -0.008 (0.008) | -0.001 (0.002) |
| **Parietal** |  |  |  |  |  |  |
| Superior Parietal | **-0.02 (0.004)** | -0.001 (0.007) | **-0.004 (0.001)** | **-0.013 (0.005)** | -0.013 (0.007) | **-0.006 (0.002)** |
| Inferior Parietal | **-0.042 (0.003)** | **-0.027 (0.007)** | **-0.003 (0.001)** | **-0.03 (0.005)** | **-0.039 (0.007)** | -0.004 (0.002) |
| Supramarginal | **-0.029 (0.003)** | -0.014 (0.007) | -0.002 (0.001) | **-0.029 (0.005)** | **-0.032 (0.007)** | -0.003 (0.002) |
| Postcentral | **-0.021 (0.004)** | 0.008 (0.007) | -0.001 (0.001) | **-0.019 (0.005)** | 0.004 (0.007) | -0.002 (0.002) |
| Precuneus | **-0.033 (0.003)** | **-0.02 (0.006)** | **-0.004 (0.001)** | **-0.028 (0.005)** | **-0.039 (0.007)** | **-0.007 (0.002)** |
| Posterior Cingulate | **-0.013 (0.004)** | **-0.025 (0.007)** | **-0.006 (0.001)** | 0.008 (0.005) | **-0.027 (0.007)** | -0.003 (0.002) |
| Isthmus Cingulate | **-0.02 (0.003)** | **-0.019 (0.007)** | **-0.003 (0.001)** | **-0.011 (0.004)** | **-0.024 (0.008)** | -0.002 (0.002) |
| **Temporal** |  |  |  |  |  |  |
| Superior Temporal | **-0.05 (0.003)** | **-0.021 (0.006)** | -0.002 (0.001) | **-0.049 (0.004)** | **-0.034 (0.007)** | -0.002 (0.002) |
| Middle Temporal | **-0.05 (0.003)** | **-0.028 (0.006)** | -0.002 (0.001) | **-0.026 (0.005)** | **-0.04 (0.007)** | -0.001 (0.002) |
| Inferior Temporal | **-0.044 (0.004)** | **-0.032 (0.006)** | -0.001 (0.001) | **-0.019 (0.005)** | **-0.047 (0.007)** | -0.001 (0.002) |
| Banks of the STS | **-0.039 (0.003)** | **-0.021 (0.007)** | -0.002 (0.001) | **-0.029 (0.005)** | **-0.031 (0.007)** | -0.002 (0.002) |
| Fusiform | **-0.041 (0.003)** | **-0.025 (0.006)** | **-0.006 (0.001)** | **-0.018 (0.006)** | **-0.031 (0.007)** | **-0.009 (0.002)** |
| Transverse Temporal | **-0.029 (0.003)** | -0.002 (0.007) | **-0.003 (0.001)** | **-0.022 (0.005)** | 0.003 (0.007) | **-0.004 (0.002)** |
| Entorhinal | **-0.017 (0.006)** | **-0.034 (0.007)** | -0.004 (0.002) | -0.007 (0.006) | **-0.04 (0.007)** | **-0.005 (0.002)** |
| Temporal Pole | **0.019 (0.007)** | -0.009 (0.008) | -0.005 (0.002) | 0.005 (0.006) | **-0.026 (0.007)** | -0.004 (0.002) |
| Parahippocampal | **-0.037 (0.005)** | **-0.022 (0.007)** | **-0.005 (0.002)** | **-0.012 (0.005)** | **-0.022 (0.007)** | **-0.004 (0.002)** |
| **Occipital** |  |  |  |  |  |  |
| Insula | **-0.019 (0.004)** | **-0.015 (0.006)** | -0.003 (0.001) | -0.01 (0.005) | **-0.033 (0.007)** | -0.003 (0.002) |
| Lateral Occipital | **-0.04 (0.003)** | -0.013 (0.006) | -0.001 (0.001) | **-0.033 (0.005)** | **-0.017 (0.007)** | -0.003 (0.002) |
| Lingual | **-0.035 (0.003)** | 0.006 (0.007) | **-0.004 (0.001)** | **-0.022 (0.005)** | 0.002 (0.008) | **-0.006 (0.002)** |
| Cuneus | **-0.028 (0.004)** | 0.009 (0.007) | **-0.003 (0.001)** | -0.008 (0.005) | 0.008 (0.008) | **-0.006 (0.002)** |
| Pericalcarine | **-0.022 (0.003)** | 0.013 (0.008) | **-0.004 (0.001)** | **-0.011 (0.004)** | 0.015 (0.008) | -0.003 (0.001) |
| **Subcortical** |  |  |  |  |  |  |
| Amygdala | **-0.063 (0.004)** | **-0.032 (0.007)** | **-0.005 (0.002)** |  |  |  |
| Caudate | **-0.007 (0.003)** | **-0.021 (0.007)** | -0.002 (0.001) |  |  |  |
| Hippocampus | **-0.067 (0.004)** | **-0.033 (0.006)** | **-0.005 (0.001)** |  |  |  |
| Lateral Ventricle | **0.05 (0.002)** | -0.003 (0.006) | **0.006 (0.001)** |  |  |  |
| Pallidum | **-0.026 (0.005)** | 0.007 (0.007) | 0 (0.002) |  |  |  |
| Putamen | **-0.042 (0.004)** | **-0.016 (0.007)** | **-0.003 (0.001)** |  |  |  |
| Significant effects after FDR-correction are highlighted in bold. | | | | | | |

| **Table 4.** **Effects of time and amyloid on volume/thickness in participants with CSF data** | | | | | | |
| --- | --- | --- | --- | --- | --- | --- |
|  | Volume |  |  | Thickness |  |  |
|  | Time | Aβ | Time × Aβ | Aβ | Time | Time × Aβ |
| **Frontal** |  |  |  |  |  |  |
| Superior Frontal | -0.008 (0.005) | -0.002 (0.012) | 0 (0.002) | -0.008 (0.007) | 0.004 (0.014) | -0.001 (0.003) |
| Rostral Middle Frontal | **-0.039 (0.004)** | 0.004 (0.012) | -0.002 (0.002) | **-0.029 (0.007)** | 0.015 (0.014) | 0.002 (0.003) |
| Caudal Middle Frontal | -0.008 (0.005) | 0 (0.014) | 0 (0.002) | -0.007 (0.007) | 0.019 (0.014) | 0 (0.003) |
| Pars Opercularis | **-0.038 (0.004)** | -0.008 (0.015) | -0.001 (0.002) | **-0.036 (0.006)** | 0.011 (0.014) | 0 (0.002) |
| Pars Orbitalis | **-0.045 (0.005)** | -0.006 (0.014) | -0.003 (0.002) | **-0.019 (0.007)** | 0.012 (0.014) | -0.003 (0.003) |
| Pars Triangularis | **-0.04 (0.004)** | 0.001 (0.014) | -0.002 (0.001) | **-0.033 (0.007)** | 0.02 (0.014) | -0.003 (0.003) |
| Medial Orbitofrontal | **-0.035 (0.006)** | 0.005 (0.012) | -0.003 (0.002) | -0.008 (0.009) | 0.004 (0.014) | -0.004 (0.003) |
| Lateral Orbitofrontal | **-0.052 (0.006)** | 0.002 (0.011) | -0.003 (0.002) | **-0.031 (0.007)** | 0 (0.015) | 0.001 (0.003) |
| Precentral | -0.012 (0.006) | 0.019 (0.013) | -0.001 (0.002) | -0.015 (0.009) | 0.017 (0.013) | -0.001 (0.003) |
| Paracentral | 0.007 (0.006) | -0.003 (0.014) | 0 (0.002) | -0.002 (0.009) | 0.005 (0.014) | -0.001 (0.003) |
| Frontal Pole | **-0.035 (0.009)** | 0.006 (0.015) | 0.007 (0.004) | **-0.04 (0.008)** | -0.014 (0.014) | 0.004 (0.003) |
| Rostral Anterior Cingulate | **-0.019 (0.005)** | -0.015 (0.013) | 0 (0.002) | **0.015 (0.007)** | -0.006 (0.015) | 0.001 (0.003) |
| Caudal Anterior Cingulate | -0.007 (0.004) | -0.001 (0.016) | **-0.005 (0.002)** | 0.011 (0.006) | -0.008 (0.016) | 0.002 (0.002) |
| **Parietal** |  |  |  |  |  |  |
| Superior Parietal | **-0.022 (0.006)** | 0.027 (0.013) | -0.003 (0.002) | **-0.021 (0.007)** | 0.015 (0.014) | -0.004 (0.003) |
| Inferior Parietal | **-0.054 (0.004)** | -0.005 (0.014) | -0.003 (0.002) | **-0.045 (0.006)** | 0.002 (0.013) | -0.004 (0.002) |
| Supramarginal | **-0.04 (0.005)** | 0.017 (0.013) | -0.002 (0.002) | **-0.038 (0.006)** | 0.011 (0.013) | -0.006 (0.002) |
| Postcentral | **-0.014 (0.005)** | 0.035 (0.014) | -0.001 (0.002) | **-0.022 (0.007)** | 0.036 (0.014) | -0.004 (0.003) |
| Precuneus | **-0.032 (0.004)** | 0.002 (0.012) | -0.002 (0.002) | **-0.039 (0.006)** | -0.011 (0.013) | -0.002 (0.002) |
| Posterior Cingulate | -0.004 (0.005) | -0.034 (0.015) | -0.002 (0.002) | -0.003 (0.006) | -0.025 (0.016) | 0.002 (0.002) |
| Isthmus Cingulate | **-0.02 (0.004)** | 0.004 (0.013) | -0.002 (0.002) | **-0.019 (0.006)** | -0.01 (0.016) | 0 (0.002) |
| **Temporal** |  |  |  |  |  |  |
| Superior Temporal | **-0.055 (0.004)** | -0.005 (0.013) | -0.003 (0.001) | **-0.05 (0.006)** | 0.007 (0.013) | -0.005 (0.002) |
| Middle Temporal | **-0.058 (0.004)** | -0.016 (0.013) | **-0.005 (0.001)** | **-0.033 (0.006)** | -0.019 (0.013) | -0.004 (0.002) |
| Inferior Temporal | **-0.057 (0.004)** | -0.006 (0.012) | -0.004 (0.002) | **-0.029 (0.006)** | -0.017 (0.014) | -0.004 (0.002) |
| Banks of the STS | **-0.048 (0.005)** | -0.017 (0.014) | -0.004 (0.002) | **-0.044 (0.006)** | -0.002 (0.014) | -0.004 (0.002) |
| Fusiform | **-0.048 (0.005)** | -0.005 (0.012) | **-0.007 (0.002)** | **-0.019 (0.007)** | 0.008 (0.014) | -0.008 (0.003) |
| Transverse Temporal | **-0.031 (0.005)** | 0.003 (0.016) | -0.005 (0.002) | **-0.027 (0.007)** | 0.04 (0.015) | -0.006 (0.003) |
| Entorhinal | -0.013 (0.008) | -0.038 (0.015) | -0.003 (0.003) | 0.01 (0.009) | -0.044 (0.016) | -0.005 (0.003) |
| Temporal Pole | 0.007 (0.011) | -0.015 (0.015) | -0.007 (0.004) | 0.006 (0.01) | -0.023 (0.015) | -0.009 (0.004) |
| Parahippocampal | **-0.036 (0.007)** | 0.006 (0.015) | -0.006 (0.003) | **-0.017 (0.006)** | -0.005 (0.015) | -0.001 (0.002) |
| **Occipital** |  |  |  |  |  |  |
| Insula | **-0.028 (0.006)** | -0.013 (0.012) | -0.002 (0.002) | 0 (0.008) | -0.036 (0.015) | -0.004 (0.003) |
| Lateral Occipital | **-0.06 (0.004)** | 0.022 (0.014) | -0.002 (0.002) | **-0.057 (0.007)** | 0.022 (0.015) | -0.002 (0.003) |
| Lingual | **-0.044 (0.004)** | 0.031 (0.016) | -0.002 (0.001) | **-0.021 (0.007)** | 0.042 (0.016) | -0.002 (0.003) |
| Cuneus | **-0.031 (0.005)** | 0.046 (0.016) | -0.002 (0.002) | -0.014 (0.007) | 0.045 (0.016) | -0.004 (0.003) |
| Pericalcarine | **-0.033 (0.005)** | 0.051 (0.017) | -0.002 (0.002) | **-0.02 (0.007)** | 0.044 (0.017) | -0.004 (0.003) |
| **Subcortical** |  |  |  |  |  |  |
| Amygdala | **-0.073 (0.007)** | -0.017 (0.014) | -0.003 (0.003) |  |  |  |
| Caudate | -0.004 (0.004) | -0.02 (0.015) | 0.001 (0.002) |  |  |  |
| Hippocampus | **-0.075 (0.005)** | -0.02 (0.012) | **-0.008 (0.002)** |  |  |  |
| Lateral Ventricle | **0.049 (0.003)** | -0.019 (0.011) | **0.005 (0.001)** |  |  |  |
| Pallidum | **-0.044 (0.007)** | 0.016 (0.013) | -0.001 (0.003) |  |  |  |
| Putamen | **-0.044 (0.006)** | 0.021 (0.013) | 0.002 (0.002) |  |  |  |
| Significant effects after FDR-correction are highlighted in bold. | | | | | | |

**Figure 1. Hippocampal volume before and after harmonization with NeuroComBat**

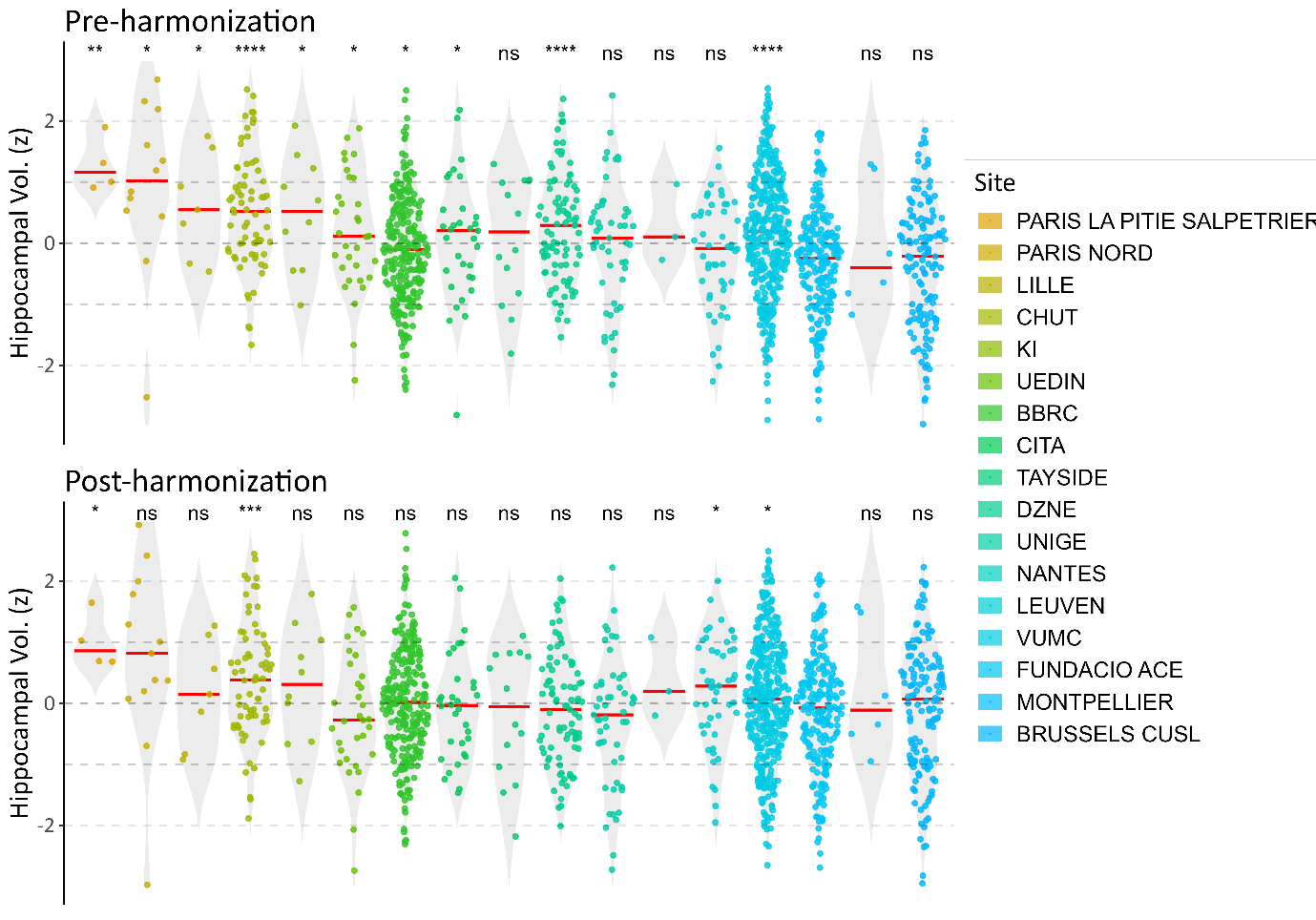
 Bilateral z-scaled hippocampal volume before and after harmonization with NeuroComBat. Differences between sites were tested with two-sample independent t-tests using ACE ALZHEIMER CENTER BARCELONA as a reference group.

**Figure 2. Distribution of Participants in Amyloid Groups**

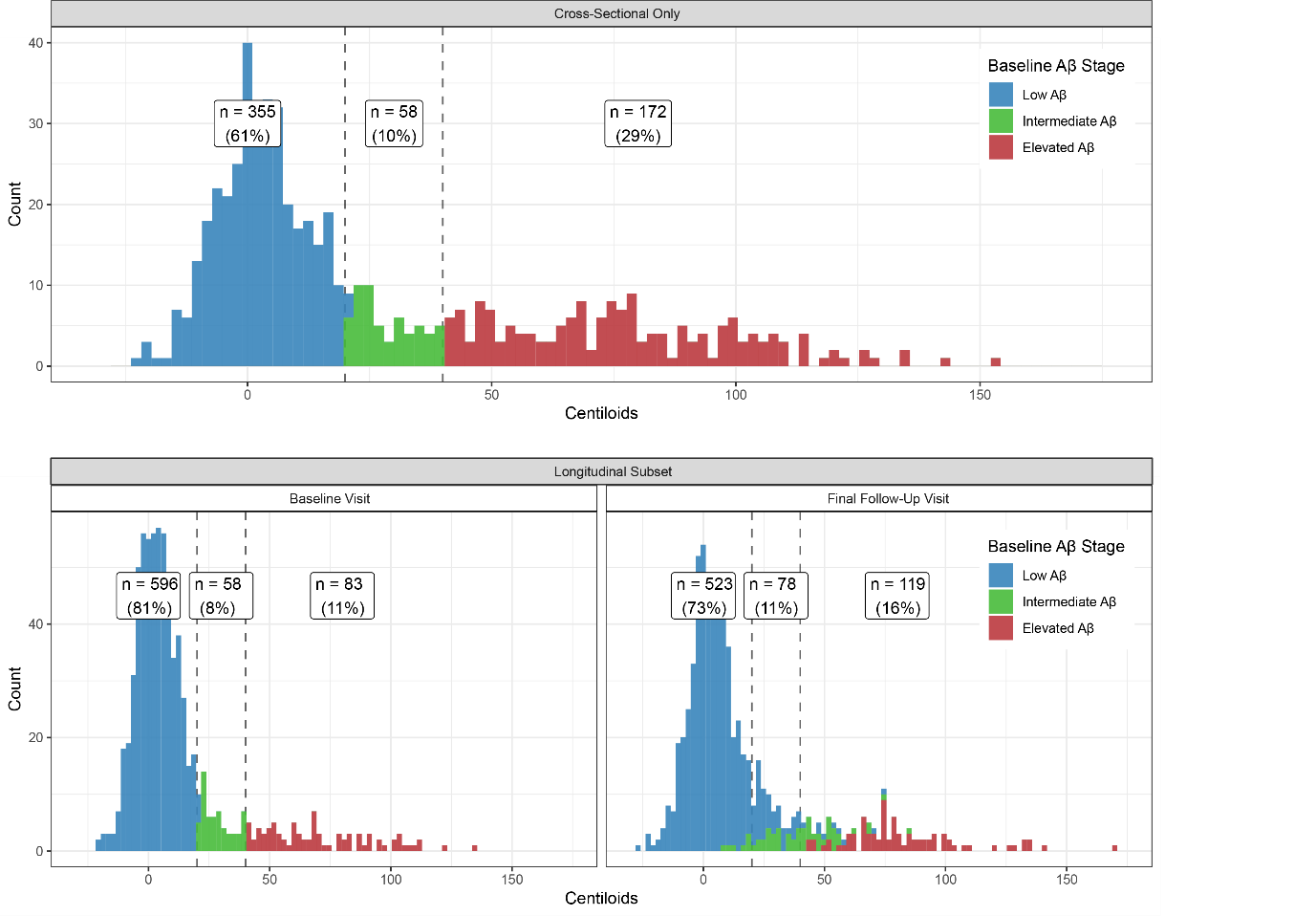

Across 585 (44.25% of all participants) participants with only one assessment at baseline (“Cross-Sectional Only”), 61% belonged to the low Aβ group, 10% to the intermediate Aβ group and 29% to the elevated Aβ group. The longitudinal subset of participants was comprised of 737 participants (55.75%), of which 81% were low Aβ, 8% intermediate Aβ and 11% baseline, which slightly shifted towards more intermediate (11%) and elevated Aβ (16%) upon follow-up.

**Figure 3. Main results replicated across amyloid stages**

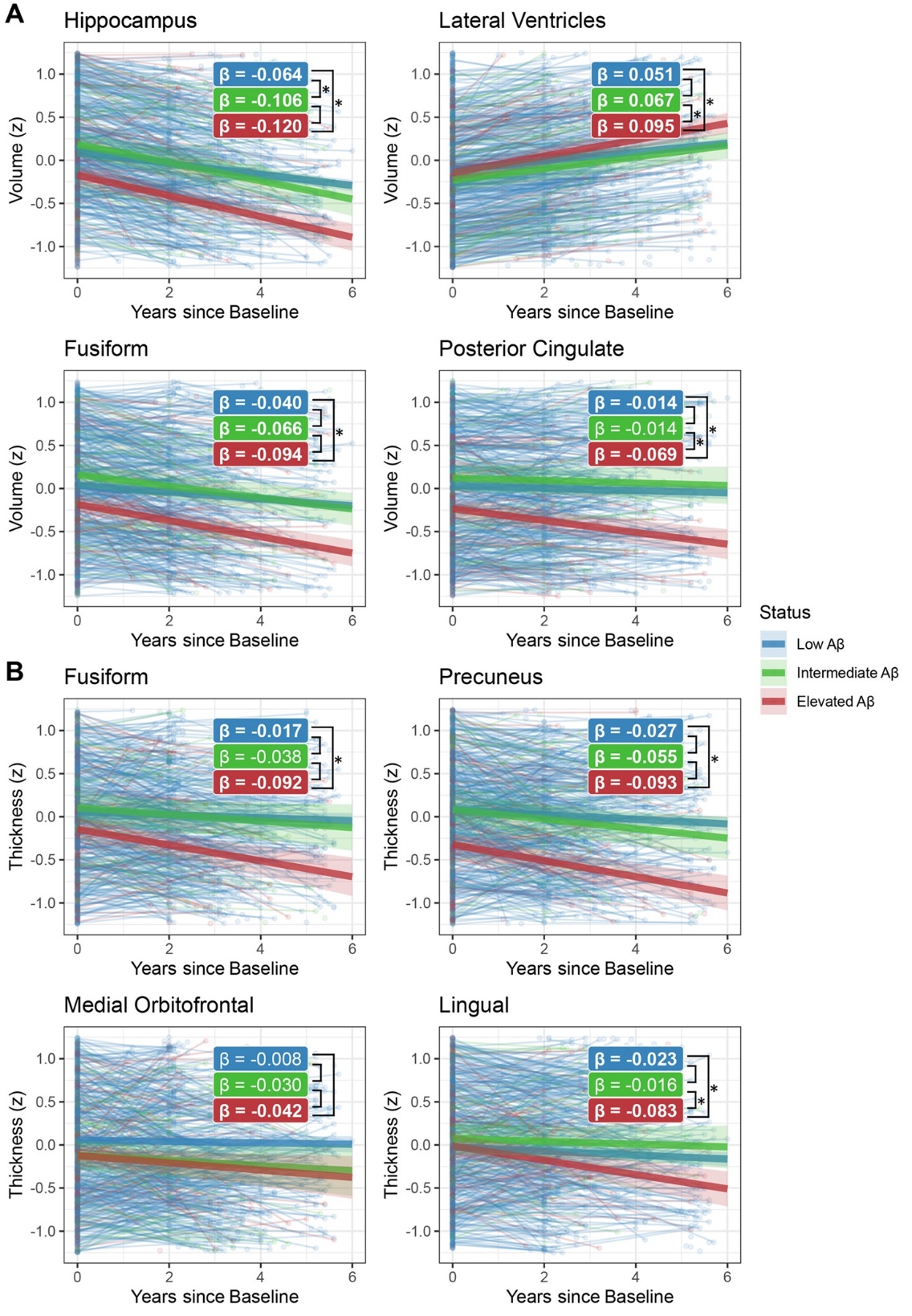

Aβ stage-wise slopes (interaction amyloid stage * time) for the four most sensitive regions for (A) volume (hippocampus, lateral ventricle, fusiform, posterior cingulate) and (B) thickness (fusiform, precuneus, medial orbitofrontal, lingual) are displayed. Significant slopes are highlighted in bold, while significant pairwise contrasts are denoted with asterisks.

**Figure 4. Longitudinal-only models and model fit improvement with time-varying Aβ**

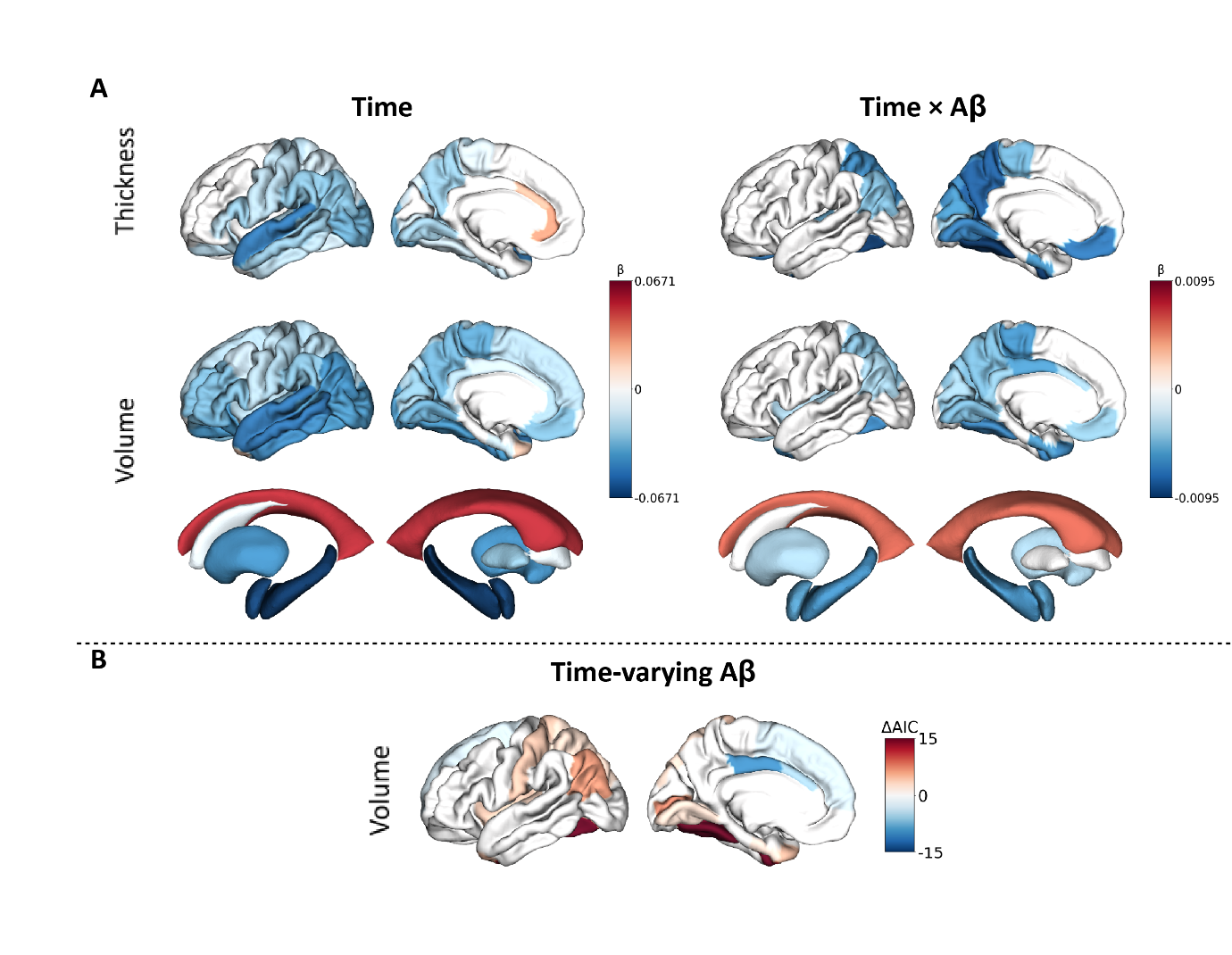

Cortical mid-surface projections of (A) FDR-thresholded LME main effect of time and interaction effect of time and Aβ on cortical thickness, cortical volume and subcortical volume; and (B) FDR-thresholded significant differences in model fit, assessed with the Akaike Information Criterion (AIC), between time-invariant baseline Aβ and time-varying Aβ. Negative AIC values indicate better model fit for time-varying Aβ. Cortical mid-surface projections show the left lateral and left medial respectively, representative of averaged bilateral effects.
